# Multi-ancestry GWAS for venous thromboembolism identifies novel loci followed by experimental validation in zebrafish

**DOI:** 10.1101/2022.06.21.22276721

**Authors:** Brooke N. Wolford, Yakun Zhao, Ida Surakka, Kuan-Han H. Wu, Xinge Yu, Catherine E. Richter, Laxmi Bhatta, Ben Brumpton, Karl Desch, Florian Thibord, Derek Klarin, Andrew D. Johnson, David-Alexandre Trégouët, Scott M. Damrauer, Nicholas L. Smith, INVENT, MVP, Valeria Lo Faro, Kristin Tsuo, Global Biobank Meta-analysis Initiative (GBMI), Mark Daly, Ben Neale, Wei Zhou, Jordan A. Shavit, Cristen. J. Willer

**Affiliations:** Department of Computational Medicine and Bioinformatics, University of Michigan, Ann Arbor, Michigan, USA; Department of Pediatrics, University of Michigan, Ann Arbor, Michigan, USA; Population Sciences Branch, Division of Intramural Research, National Heart, Lung and Blood Institute, 73 Mt. Wayte, Suite #2, Framingham, MA, 01702, USA; Division of Vascular Surgery, Stanford University School of Medicine, Palo Alto, CA, 94305, USA; Department of Internal Medicine, University of Michigan, Ann Arbor, Michigan, USA; Division of Cardiovascular Medicine, Department of Internal Medicine, University of Michigan, Ann Arbor, Michigan, USA; K.G. Jebsen Center for Genetic Epidemiology, Department of Public Health and Nursing, NTNU, Norwegian University of Science and Technology, Trondheim, 7030, Norway; HUNT Research Centre, Department of Public Health and Nursing, NTNU, Norwegian University of Science and Technology, Levanger, 7600 Norway; Clinic of Medicine, St. Olavs Hospital, Trondheim University Hospital, Trondheim, 7030, Norway; Univ. Bordeaux, Inserm, Bordeaux Population Health Research Center, UMR 1219, F-33000 Bordeaux, France; Corporal Michael Crescenz VA Medical Center, Philadelphia PA, USA; Department of Surgery, Perelman School of Medicine, University of Pennsylvania, Philadelphia PA, USA; Department of Genetics, Perelman School of Medicine, University of Pennsylvania, Philadelphia PA, USA; Cardiovascular Health Research Unit, Departments of Biostatistics and Medicine, University of Washington; University of Groningen, UMCG, Department of Ophthalmology, Groningen, the Netherlands; Department of Clinical Genetics, Amsterdam University Medical Center (AMC), Amsterdam, the Netherlands; Department of Immunology, Genetics and Pathology, Science for Life Laboratory, Uppsala University, Uppsala, Sweden; Analytic and Translational Genetics Unit, Department of Medicine, Massachusetts General Hospital, Boston, Massachusetts, USA; Stanley Center for Psychiatric Research, Broad Institute of MIT and Harvard, Cambridge, Massachusetts, USA; Program in Medical and Population Genetics, Broad Institute of Harvard and MIT, Cambridge, Massachusetts, USA; Institute for Molecular Medicine Finland, University of Helsinki, Helsinki, Finland; Department of Human Genetics, University of Michigan, Ann Arbor, Michigan, USA; University of Toledo, College of Medicine and Life Sciences

**Keywords:** Venous thromboembolism, GWAS, meta-analysis, mendelian randomization, crispant

## Abstract

Genome wide association study (GWAS) results for Venous Thromboembolism (VTE) across 9 international cohorts of the Global Biobank Meta-analysis Initiative (GBMI), with representation across six ancestry groups (cases=27,987, controls=1,035,290), were combined using inverse-variance weighted meta-analysis. This multi-ancestry GWAS resulted in 38 genome-wide significant loci, 9 of which are potentially novel. For each autosomal locus we performed gene prioritization using seven independent, yet converging, lines of evidence. Through prioritization we identified genes known for VTE (e.g., *F5, F11, VWF*), genes known to modify blood coagulation (e.g., *STAB2*), and genes without known coagulation mechanisms from functional studies (e.g., *PLCG2, TC2N*). We evaluated the function of six prioritized genes, including *F7* as a positive control, using laser mediated endothelial injury to induce thrombosis in zebrafish. We used CRISPR/Cas9 to knock down these potentially causal genes and measured time to occlusion after laser injury. From this assay we have supportive evidence for a role of *RASIP1* and *TC2N* in the modification of human VTE, and suggestive evidence for *STAB2* and *TSPAN15*. This study expands the currently identified genomic architecture of VTE through biobank-based multi-ancestry GWAS, *in silico* candidate gene predictions, and *in vivo* functional follow-up of novel candidate genes.

## INTRODUCTION

Deep vein thrombosis and pulmonary embolism, collectively referred to as venous thromboembolism (VTE), are disorders characterized by the pathologic formation of thrombi in deep veins that may embolize to the pulmonary circulation. VTE is a common cause of morbidity and mortality affecting >900,000 individuals per year in the United States(CDC, 2020a, 2020b; Heit, 2015; Henke et al., 2020). As a complex trait, VTE risk is influenced by an array of well described environmental factors as well as genetics. Heritability studies suggest that between 30-40% of VTE risk is due to genetic factors(PMID: 34913365). Before genotyping technology allowed a genome-wide assessment of common variants, many common polymorphisms were implicated in VTE risk, but only two were validated by initial European ancestry GWAS studies. Those were the ABO blood group(Souto et al., 2000) and a variant in the procoagulant gene *F5* (Factor V Leiden)(Shavit and Ginsburg, 2013). Sixteen previous large genome-wide association studies (GWAS) and exome sequencing studies have identified common and rare variants from dozens of loci associated with risk for VTE. References are provided in **Supplementary Table 1**, and a “gold standard” list of coagulation and platelet genes(Downes et al., 2019) is in **Supplementary Table 2**.

The strongest signals observed in GWAS are those associated with loci with known roles in the hemostatic system such as gain of function variants in procoagulant genes (*F2, F5, F11, FGG(Klarin et al., 2019)*) or missense variants p.Ser219Gly and p.Arg113Cys in anticoagulant gene *PROCR(Dennis et al., 2012; Manderstedt et al., 2022)*. As the list of VTE risk variants continues to grow and includes loci with no previously described genes involved in thrombosis, so does the need for functional analyses of these variants. Zebrafish are a vertebrate model which display a high degree of conservation with the human genome(Howe et al., 2013), including the coagulation cascade(Kretz et al., 2015). Their high fecundity, optical transparency, and external development make them amenable to rapid analysis of GWAS signals in an *in vivo* model(Liu et al., 2013). We have previously shown that we can evaluate normal and pathologic hemostasis, as well as thrombosis, in zebrafish embryos and larvae using genome editing(Liu et al., 2014; Rost et al., 2016).

Here we expand the list of loci associated with VTE through a multi-ancestry meta-analysis of GWAS from biobanks of the Global Biobank Meta-analysis Initiative (GBMI). Using genetic associations at known and potentially novel loci, we perform integrative bioinformatics-driven gene prioritization and subsequent functional analyses of seven candidate genes in zebrafish assessed for thrombosis.

## RESULTS

### Multi-ancestry meta-analysis yields 9 potentially novel loci

We performed single variant association analyses in nine biobanks with diverse ancestries (Supplementary Figure 1), followed by meta-analysis, to look for VTE-associated loci. The prevalence of VTE was much higher in biobanks with hospital-based recruitment (e.g., UCLA) versus the <2% observed in population based biobanks (e.g., ESTBB), highlighting the important role that sampling strategy plays in identifying cases. Overall, top loci had higher effect sizes in the population-based biobank meta-analysis than the hospital-based biobank meta-analysis **(Supplementary Figure 2)**. We identified 38 genome-wide significant loci (**Table 1, Supplementary Table 3**). Of these, 31 lead SNPs are within 500 kb of a variant previously reported in GWAS or sequencing studies, resulting in 9 potentially novel associations. Three variants (at *ABO, FGG* and *PROCR* loci) had nominally significant heterogeneity across ancestries (Cochran’s Q p-value < 0.05), but were not considered significant after Bonferroni correction for multiple testing (**Supplementary Figure 3**). We identified a potentially novel locus (rs112106699 near *DHRS3*) that is rare in European ancestries (gnomAD v2.1.1 Non Finnish European allele frequency(AF)=0.06%) but driven by higher frequency in African ancestry cohorts (gnomAD African/African American AF=9.0%), highlighting the importance of multi-ancestry meta-analysis (**Supplementary Figure 4**). We also identified common 2bp indel and 66bp structural variants that were associated with VTE (rs10559566 near *CARMIL1/SCGN* and rs1459062246 near *HOXB2*) and two additional rare variants (each < 0.5% frequency) with large effect sizes (OR > 2.1; rs562281690 near *EPHA3* and rs115924439), highlighting the importance of inclusion of common and rare indels, structural and single variants in GWAS.

**Table 1.**
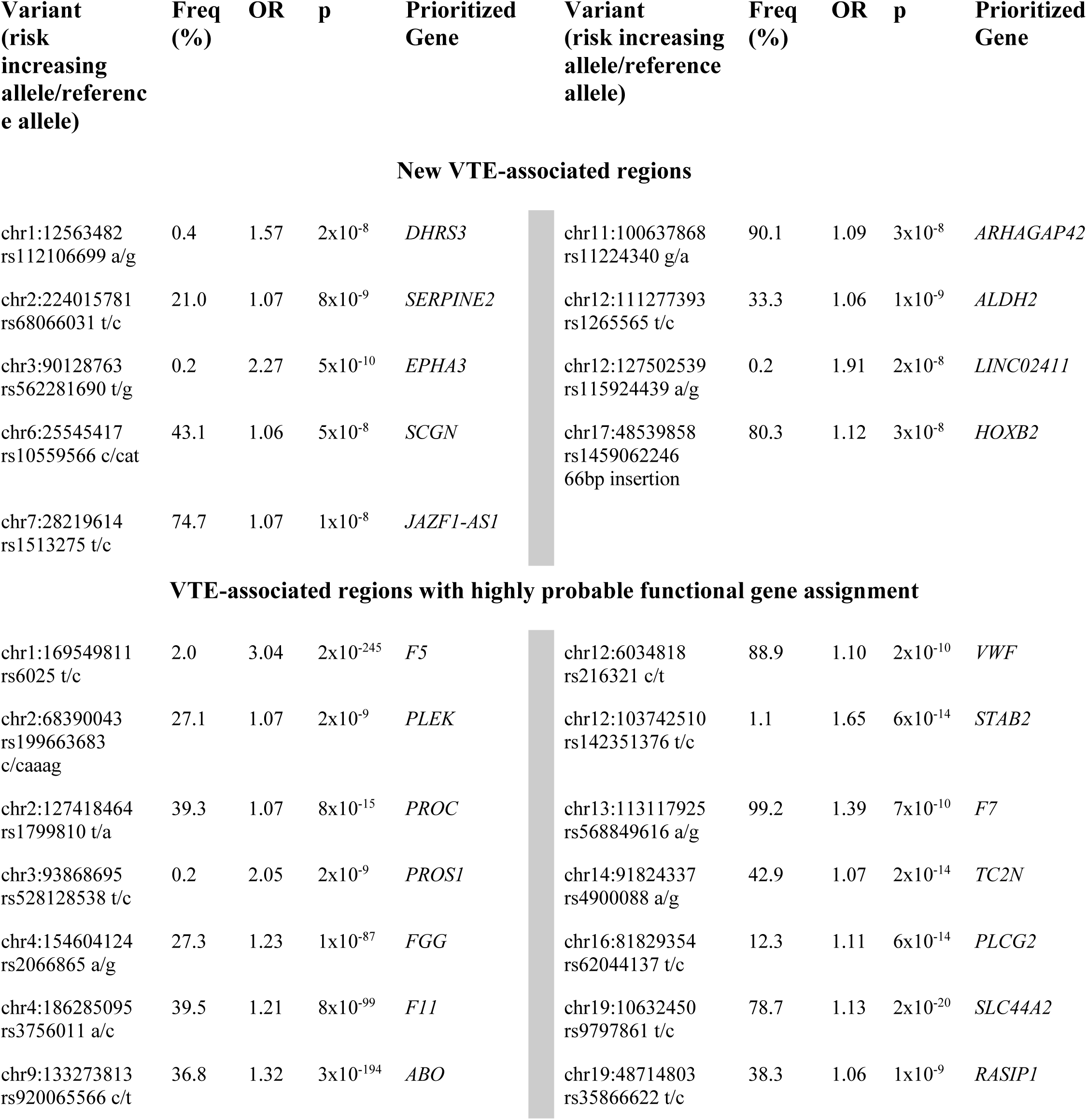
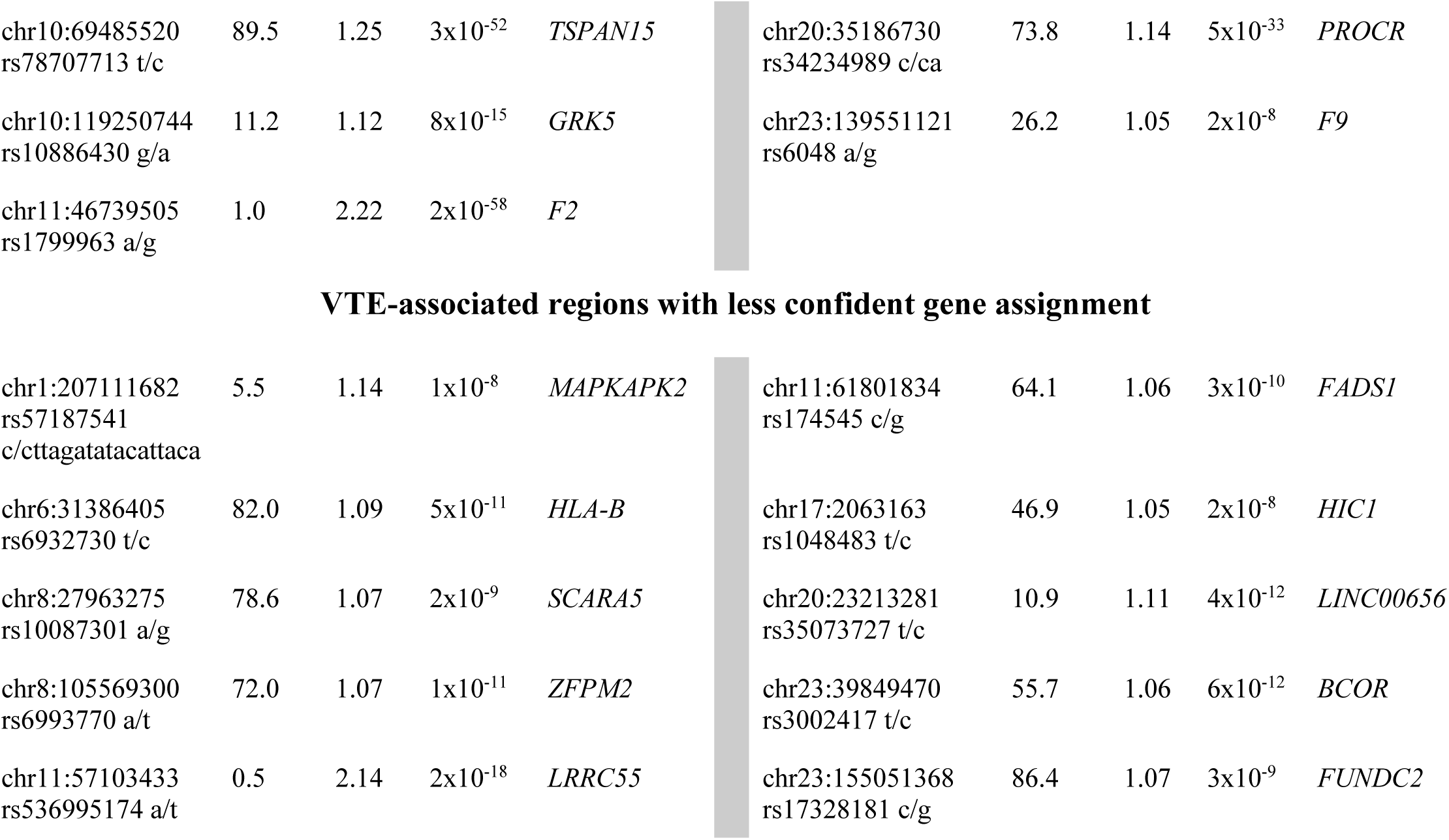
Genome-wide significant VTE loci. Of the 38 genome-wide significant loci, 9 are potentially novel. Of the regions previously identified by GWAS, 19 genes have highly probable functional gene assignment through integrative gene prioritization and literature. Finally, 10 loci have less confident functional gene assignment. These genes had 3 or fewer lines of evidence in the integrative prioritization and were not known from previous function studies.

We performed replication with INVENT+MVP VTE association summary statistics (N=42,032 discovery cases)(Thibord et al., 2022). All 38 associated loci were tested in the replication dataset (rs187506383 was used as a proxy for rs536995174 [linkage disequilibrium = r^2^>0.8], the other 37 index variants were available). 35 of the 38 lead SNPs from the discovery meta-analysis were nominally significant in the replication, with 30 significant at a Bonferroni threshold of 1.4×10^−3^. This is significantly more replicated loci than we would expect by chance (binomial test p-value 0.00024). Five of the 30 replicated lead SNPs are in loci considered to be potentially novel. All of the variants showed consistent direction of effect between discovery and replication cohorts, with a Pearson’s correlation of discovery and replication effect sizes of 0.93 (95% CI 0.87, 0.96) (**Supplementary Figure 5**). Due to a potential overlap of individuals between eMERGE samples in INVENT and BioVU samples in GBMI, we also compared INVENT summary statistics to GBMI summary statistics without BioVU as a sensitivity analysis (**Supplementary Figure 5**) and observed similar results (Pearson’s correlation 0.92, 95% CI 0.84-0.96). Additionally, when meta-analyzing the replication summary statistics with leave-BioVU-out summary statistics from GBMI, 35 loci still show a combined P-value of <5×10^−8^ (**Supplementary Table 4**). Furthermore, the broad agreement between replication and discovery datasets gives us confidence in the phenotype used for GBMI, despite inclusion of nurse-verified, self reported cases in INVENT+MVP but not in GBMI.

### Integrative gene prioritization nominates likely causal genes

For each autosomal locus (N=36) we performed gene prioritization using seven independent lines of evidence. For each locus, the functional gene we identified i) has a missense variant within the 95% credible variant set (8 loci), ii) is the closest gene within 1 kb (25 loci), ii) is the only gene prioritized by DEPICT (8 loci), iii) is a gene within the top 10% of PoPS scores (17 loci), iv) is an expression quantitative trait loci (eQTLs; 4 loci), v) found in ClinVar for related phenotypes (5 loci), or vi) prioritized via colocalization and proteome-wide Mendelian randomization (PWMR,15 genes at 23 loci, **Supplementary Table 3)**. Notably, eQTL and ClinVar variants must be the lead SNP at that locus.

By summing over the number of lines of evidence supporting a given gene, we prioritized at least one gene at 31 of the 36 loci—a single likely causal gene for 28 of the 36 loci and two prioritized genes for three loci (**Supplementary Table 3**). The genes with the most evidence (5 of 7) as likely causal genes are *F5, PLEK*, and *PROS1* (**Figure 1**). Similarly, genes that had four lines of evidence supporting a likely causal gene are *PROC, FGG, F11, F2, VWF*, and *PLCG2*. Notably, all genes with four or five lines of evidence were previously reported by GWAS and all except for *PLCG2* were previously known by functional studies. Of the 9 potentially novel loci from our GWAS, we prioritized one or two likely causal genes for 7 of these loci, including some with three lines of evidence (*SERPINE2, JAZF1-AS1*, and *ALDH2*), two lines of evidence (*HOXB2, SCGN*) and only 1 evidence (*DHRS3, EPHA3*). At the three loci on the X-chromosome, although the full complement of gene prioritization approaches weren’t available due to bioinformatics limitations, the three genes closest to the index variants were likely causal genes. On the X chromosome, common variant p.Thr194Ala in *F9* known as F9 Malmö (rs6048)(Bezemer et al., 2009) was associated in our dataset, as well as a variant ∼200kb upstream of *BCOR* (rs3002417). An intronic variant of *FUNDC2* (rs17328181) previously nominally associated with VTE (p-value=2×10^−7^)(Lindström et al., 2019) was also associated. *FUNDC2* is known to be involved in platelet survival(Ma et al., 2019) and is ∼29kb from the 5’ end of *F8*, a gene in which genetic mutations cause Factor VIII deficiency or hemophilia A and thrombosis(Kamphuisen et al., 2001).

**Figure 1.**
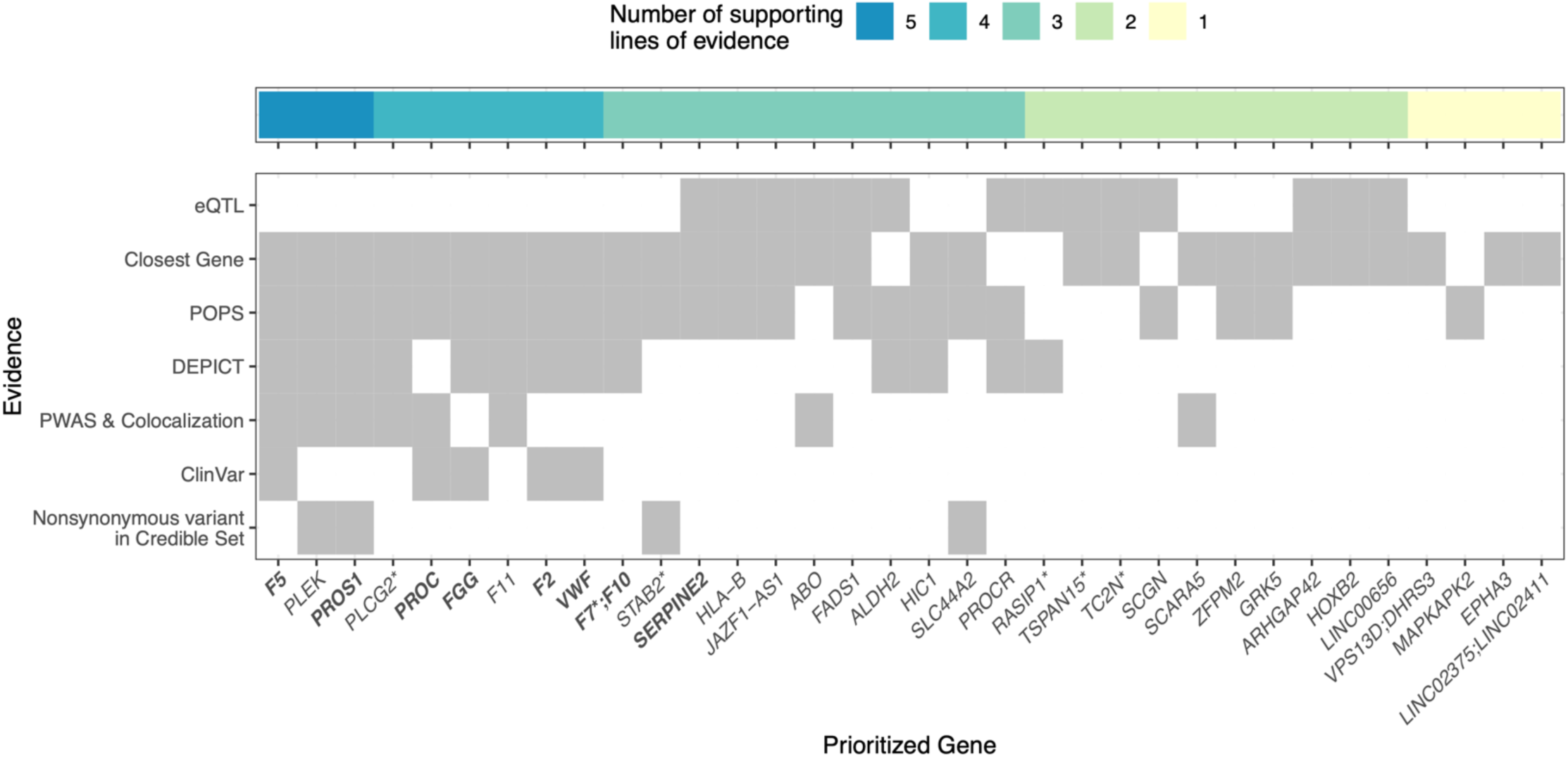
Integrative gene prioritization. Autosomal genome wide significant loci labeled by prioritized gene (x-axis) with shading for each line of evidence used in the bioinformatics-driven prioritization scheme (y-axis). Genes in bold were on the gold standard list (**Supplementary Table 2**). For *VPS13D;DHRS3, F7;F10*, and *LINC02375;LINC02411* the genes had equal number of supporting lines of evidence. *LINC00656* is also known as *RP4-737E23*.*2*. rs536995174 had one line of evidence each for *SERPING1, SLC43A3, SLC43A1, F2, OR5AK4P, LRRC55* and has been excluded from this visualization. Genes with asterisks were selected for follow up with a functional assay in zebrafish.

Using our integrative gene prioritization approach we identified eleven genes known to be involved in blood clotting (*F2, F5, F9, F7, F10, F11, FGG, PROC, PROCR, PROS1, VWF*), including genes known to regulate blood clotting factors from functional analyses but not previously identified by GWAS (e.g., *PROS1, STAB2, SERPINE2*), and genes without known mechanisms (e.g., *TC2N, PLEK*). Using Enrichr for the 38 prioritized candidate genes (one per locus), we identified significantly enriched gene sets including; KEGG ‘complement and coagulation cascades’ (**Supplementary Table 5**, 10 genes highlighted; enrichment p=4×10^−^ 14), Gene Ontology Biological Process of ‘negative regulation of blood coagulation’ (**Supplementary Table 6**, 6 genes highlighted; enrichment p=6×10^−8^) and Gene Ontology Molecular Function ‘serine-type endopeptidase activity’ (**Supplementary Table 7**, 5 genes highlighted; enrichment p=1×10-4).

### In vivo functional analyses in zebrafish provide evidence for causal role of *RASIP1* and *TC2N*

Gene prioritization has shown evidence that a number of known coagulation factors contribute to VTE. Our previous studies validated several known coagulation factors using the genome edited zebrafish models of hemostasis and thrombosis: *F2* (Grzegorski et al., 2020) *F5(Weyand et al., 2019), F10(Hu et al., 2017), PROS1(Ku et al., 2020)*, and *PROC(Ku et al., 2020)*. Using this model, we evaluated 6 prioritized genes (*F7, RASIP1, TC2N, STAB2, TSPAN15*, and *PLCG2)* from regions that demonstrate conservation of synteny with the zebrafish genome. We chose genes across the spectrum of gene prioritization to see if prioritized genes with more lines of evidence are more likely to be functional. *PLCG2* had the most lines of supporting evidence of the genes that are novel when considering functional studies. From genes with three lines of evidence, *F7* was previously used as a positive control in this assay and *STAB2* has a missense variant in the 95% credible set. When selecting the final genes, we selected those with the most significant p-values amongst the genes with two lines of evidence.

CRISPR/Cas9 was used to create mosaic knockdown embryos (Burger et al., 2016) (“crispants”) for genotype-blinded evaluation of time to occlusion after induced endothelial laser injury at 3 days post fertilization. Based on increased time to occlusion in knockdown versus uninjected controls, and after accounting for Bonferroni multiple testing correction, we have supportive evidence for a role of *RASIP1* (WRST p-value 2.8e-15) and *TC2N* (WRST p-value 8.1e-4) in the modification of human VTE (**Figure 2**). *STAB2 and PLCG2* were nominally significant (p-value < 0.05). To increase power, we also pooled the non-injected controls from multiple experiments and compared the median time to occlusion to the median for each knockdown gene. This comparison confirmed *RASIP1* and provided suggestive evidence for *STAB2* and *TSPAN15* as involved in coagulation following endothelial injury.

**Figure 2.**
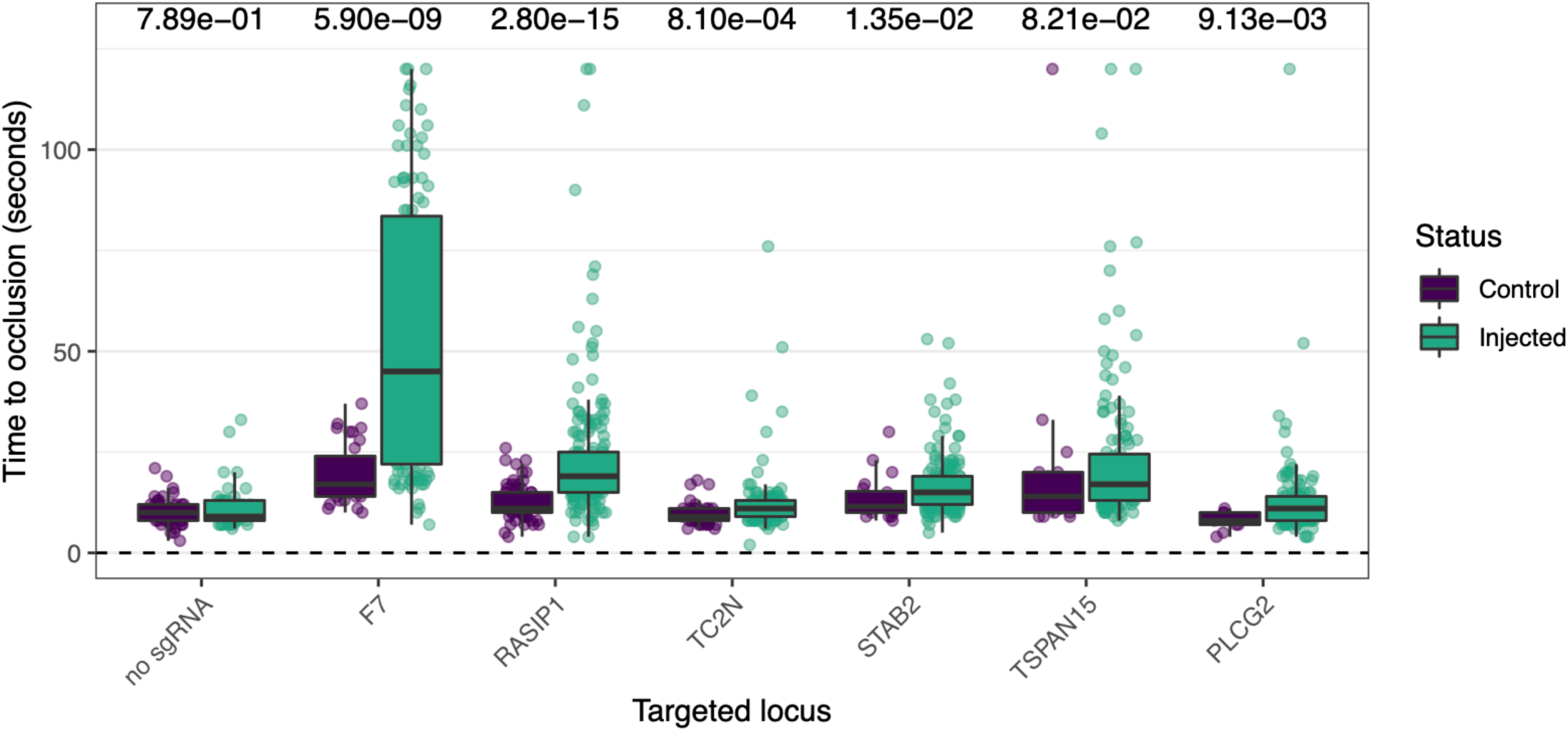
Functional evidence for causal genes in genetically modified zebrafish. P-values from Wilcoxon Rank Sum Test are listed above. The y-axis is experimental times to occlusion for control and sgRNA injected zebrafish embryos with the x-axis showing the genes targeted via CRISPR. Injections made without single guide RNA (sgRNA) serve as a negative control. Factor 7 (*F7)* serves as a positive control.

### Genetic predisposition to VTE is causally associated with COVID-19

The genetic correlation between COVID-19 severity and risk factors, biomarkers, and complex diseases was previously quantified(COVID-19 Host Genetics Initiative, 2021). However, VTE was not included in the analysis. Unusual clotting and bleeding has been observed in COVID-19 patients(Al-Samkari et al., 2020) with incidence of VTE around 20% in hospitalized patients(Middeldorp et al., 2020) and 15% prevalence of COVID-19 related VTE(Tan et al., 2021). We estimated that individuals with VTE had an 8% increased risk of hospitalization due to COVID-19 (IVW odds ratio 1.08; 95% CI 1.01–1.17, p-value 0.034, **Table 2**). Sensitivity analysis such as weighted median, penalized weighted median, and weighted mode were performed and presented in **Table 2** and **Figure 3**. Other than MR Egger (OR 1.15, 95% CI 1.02-1.29, p-value=0.041), the estimates were slightly attenuated in these sensitivity analyses compared to the main analysis. Strong evidence of heterogeneity (Q statistic=59.97, p-value <0.001) was observed, which suggests the presence of some invalid instruments. Furthermore, we repeated the main analysis using MR-PRESSO, which excluded three SNPs being horizontal pleiotropic outlier variants. Causal estimates observed in MR-PRESSO (outlier corrected odds ratio 1.05; 95% CIs 1.01–1.09, p-value 0.024) were similar to the main analysis, implicating increased genetic risk for VTE as a risk factor for hospitalization due to COVID-19.

**Table 2.** Causal association of VTE with COVID19 hospitalization using two-sample Mendelian randomization. Table of two-sample mendelian randomization (MR) estimates for the causal association of VTE with COVID19 hospitalization. The odds ratios (ORs) presented explained the increased risk of COVID19 hospitalization for the people with VTE. In addition to OR for VTE from IVW methods for two-sample MR analysis, ORs from 5 other methods as sensitivity analysis are presented.

| **Methods** | **No. of SNPs** | **OR** | **95% CI** | **p-value** |
| --- | --- | --- | --- | --- |
| **IVW** | 18 | 1.08 | 1.01 – 1.17 | 0.034 |
| **MR-Egger** | 18 | 1.15 | 1.02 – 1.29 | 0.040 |
| **Weighted median** | 18 | 1.04 | 0.98 – 1.11 | 0.185 |
| **Penalized weighted median** | 18 | 1.03 | 0.97 – 1.10 | 0.292 |
| **Weighted mode** | 18 | 1.03 | 0.97 – 1.09 | 0.345 |
| **MR-PRESSO** | 15 | 1.05 | 1.01 – 1.09 | 0.024 |

**Figure 3:**
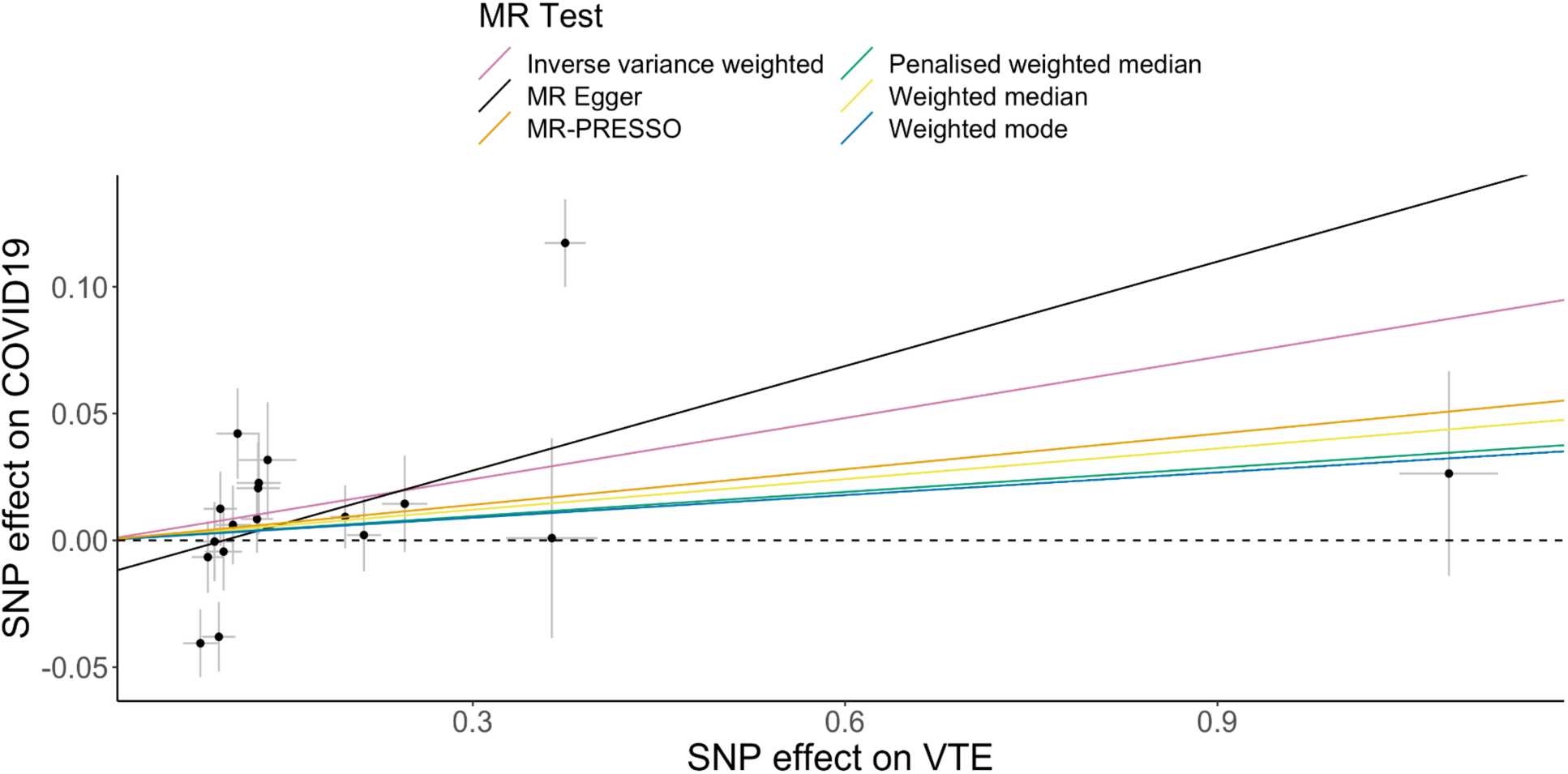
Scatter plot for the causal association of VTE with COVID19 hospitalization using two-sample Mendelian randomization. Scatterplot of genetic instruments used in two-sample mendelian randomization (MR). The effect size for the SNP association with hospitalized COVID-19 from the COVID-19 Host Genetics Initiative meta-analysis is on the y-axis, and the effect size for the association with VTE from GBMI meta-analysis in European ancestries. Each point has 95% confidence intervals for each association. In addition to slope of inverse variance weighted (IVW, pink) method for two-sample mendelian randomization analysis, slopes of five other methods as sensitivity analysis are presented.

## DISCUSSION

In this study, we perform a multi-ancestry GWAS of VTE, replicate the findings in another multi-ancestry GWAS of VTE, and identify 9 potentially novel loci. Using a bioinformatically-driven gene prioritization heuristic, we identify prioritized genes at each locus, and perform a laser-mediated injury assay in zebrafish with CRISPR knockdown of 6 putative causal genes. We also use significant SNPs as genetic instruments to test for a potential causal relationship between severe COVID-19 and VTE.

The replication of 30 of 38 lead SNPs in an independent multi-ancestry meta-analysis associated with VTE lends credibility to the GBMI’s ICD-code based meta-analysis across ancestries. While evidence for heterogeneity between studies was minimal, some spurious associations are still possible. The integrative prioritization method is similar to previous studies, but no gold standard method exists for defining the most probable causal gene. One limitation of the credible sets used for prioritization is the unreliability of the fine-mapping results from a multi-ancestry meta-analysis(Kanai et al., 2021).

We can assign a candidate causal gene to 7 of the 9 novel loci after augmenting bioinformatics-driven integrative gene prioritization with literature review (**Table 1**). The intronic variant rs68066031 on chromosome 2 is a cis-eQTL for *SERPINE2* in whole blood and is the first report of human genetic association evidence at *SERPINE2*, which encodes a serine protease inhibitor known as nexin-1 which binds heparin and inhibits thrombin(Bouton et al., 2012). Of note, a common treatment for VTE are direct oral anticoagulants (DOACs) (Elsebaie et al., 2019) which inhibit thrombin like *SERPINE2*. On chromosome 6, the intronic variant rs10559566 is in *CARMIL1*, a gene previously associated with platelet counts(Wei et al., 2017) and is the highest prioritized gene via PoPS, however the strongest eQTLs for this variant point to *SCGN*. The location of the lead SNP and eQTLs point to *JAZF1-AS* as the likely causal gene of intronic noncoding RNA variant rs1513275 on chromosome 7, although *JAZF1* is known to be associated with type 2 diabetes(Zeggini et al., 2008). On chromosome 11, the intergenic variant rs11224340 is both an eQTL in tibial arterial tissue for *ARHGAP42* and 46 kb away. *ARHGAP42* been previously associated blood pressure (Kato et al., 2015) while *CNTN5*, 278 kb away, has been associated with platelet count(Sakaue et al., 2021), white blood cell count(Sakaue et al., 2021), and red blood cell distribution width(Astle et al., 2016). On chromosome 12 an intronic variant rs1265565, is in *CUX2*, which has previously been associated with developmental and epileptic encephalopathy(Chatron et al., 2018). 55 kb away, *SH2B3* has been associated with thrombocythemia, a disorder of excessive blood platelet production (ClinVar; Oh et al., 2010). *SH2B3* and *ALDH2* (416 kb away) are both prioritized by DEPICT and PoPS, and *ALDH2* is also a significant eQTL in arterial aorta. On chromosome 17 the insertion falls into a gene-enhancer region between *HOXB1* and *HOXB2*, and near *HOXB2-AS1*. While there are blood trait associations in the GWAS catalog, it is unclear which *HOX* gene in this region may be causal, although the associated lead variant is 2.4kb upstream and also an eQTL for *HOXB2*. The potentially novel variants on chromosome 1 (*DHRS3*), chromosome 3 (*EPHA3*), and chromosome 12 (*LINC02411)* do not have clear candidate causal genes and are assigned based on proximity.

While imperfect, the integrative gene prioritization provided a starting point for functional genomic study of VTE. Although there was not significantly different time to occlusion between knock downs and controls after accounting for multiple testing, *STAB2, TSPAN15*, and *PLCG2* remain candidates as causal genes. For *STAB2*, sequencing of 393 VTE cases and 6,114 controls identified rare, damaging variants in the gene with strong evidence for a role in modifying thrombosis risk(Desch et al., 2020). Lack of functional confirmation could be due to limited statistical power, insufficient knockdown, or a failure of the zebrafish molecular readout to replicate the *in vitro* function of these genes in humans. Currently, knock out models for *TSPAN15* and *PLCG2* are being created for testing of thrombotic activity. This assay only tests the ability to produce fibrin-rich thrombi. It is possible that these genes affect thrombosis through modification of other pathways involved in clotting, including effects on platelets or the vasculature. For example, although *RASIP1* knockdowns did significantly alter the time to occlusion, this gene is known to be a regulator of vascular integrity(Wilson et al., 2013) and therefore might also mediate VTE via mechanisms not evaluated in this assay. The additional prioritized genes from the genome-wide significant loci remain intriguing for functional follow-up in future studies.

Coagulation abnormalities have been well documented in patients with severe COVID-19(Levi et al., 2020). Furthermore, remote history (>9 years previously) of VTE is associated with increased risk of COVID-19(Anderson et al., 2021). Previous Mendelian randomization (MR) studies focused on coagulation factors specifically observed that von Willebrand Factor (VWF) is the only one with genetic causal association with the incidence of severe COVID-19(Zhou et al., 2021b) to date. Another recent study(Leong et al., 2021) using genetic instruments from a GWAS of European ancestry found no significant association between VTE and severe COVID-19. Given the previously established causality of severe COVID-19 on coagulopathies, this direction of causality rather than the opposite allowed us to identify VTE as an important risk factor that could be used for risk stratification. However, given the small effect size (1.08, 95% CI 1.01-1.17) and the rarity of VTE in the general population, there are no immediate clinical translations for this finding.

### Outlook/Conclusion

This work contributes to the understanding of genetic variation associated with VTE. We identified 38 genome wide significant loci, 30 of which were replicated, and included 9 potentially novel loci. Using an integrative prioritization approach, we identified potentially causal genes with a high concordance with known functional studies, GWAS findings, and literature. We followed up 6 genes with new functional studies in zebrafish to investigate their role in thrombosis. Finally, a better understanding of the human genetic background of VTE will lead to new insights into VTE pathophysiology. These suggest novel diagnostic and therapeutic targets for management of thrombosis in affected patients.

## Supporting information

GBMI-VTE-Supplemental-Material

GBMI-VTE-Supplemental-Tables

## Data Availability

All summary statistics will be released with the GBMI flagship paper. All functional data in the present study are available upon request. Code used is available on github.

https://github.com/bnwolford/gbmi_vte

## Acknowledgments

The authors acknowledge the biobank participants, recruitment teams and project managers of the Global Biobank Meta-analysis Initiative for providing their data for biomedical research, providing data aggregation, management, and distribution services in support of the research reported in this publication (especially Sinéad Chapman and Bethany Klunder). We acknowledge BioBank Japan (Yukinori Okada, Koichi Matsua, and Masahiro Kanai), BioMe (Ruth Loos, Judy Cho, Eimear Kenny, Michael Preuss, and Simon Lee), BioVU (Nancy Cox and Jibril Hirbo), Canadian Partnership for Tomorrow (Philip Awadalla and Marie-Julie Fave), China Kadoorie (Robin Walters, Kuang Lin, and Iona Millwood), Colorado Center for Personalized Medicine (Kathleen Barnes, Michelle Daya, and Chris Gignoux), deCODE Genetics (Kári Stefánsson and Unnur Þorsteinsdóttir), East London Genes & Health (David A van Heel, Sarah Finer, and Richard Trembath), Estonian Biobank (Andres Metspalu, Reedik Mägi, Tõnu Esko, and Priit Palta), FinnGen (Aarno Palotie, Mark Daly, Samuli Ripatti, Mitja Kurki, and Juha Karjalainen), Generation Scotland (Caroline Hayward and Riccardo Marioni), HUNT (Kristian Hveem, Cristen Willer, and Sarah Graham, Ben Brumpton, and Brooke Wolford), Lifelines (Serena Sanna and Esteban Lopera), Michigan Genomics Initiative (Sebastian Zoellner, Michael Boehnke, Lars Fritsche, and Anita Pandit), Million Veteran Program (Christopher J. O’Donnell), Netherlands Twin Register (Dl Boomsma, MG Nivard), Partners Biobank (Jordan Smoller and Yen-Chen Feng), QIMR Berghofer (Sarah Medland, Stuart McGregor, and Nathan Ingold), Taiwan Biobank (Yen-Feng Lin, Yen-Chen Feng, and Hailiang Huang), UCLA Precision Health Biobank (Ruth Johnson, Yi Ding, Alec Chiu, Bogdan Pasaniuc, and Daniel Geschwind), and UK Biobank (Konrad Karczewski and Alicia Martin).

JS was supported by R35 HL150784. D-AT was supported by the EPIDEMIOM-VT Senior Chair from the University of Bordeaux initiative of excellence (IdEX) and the GENMED Laboratory of Excellence on Medical Genomics [ANR-10-LABX-0013], a research program managed by the National Research Agency (ANR) as part of the French Investment for the Future. CJW, IS, KHW, and BNW were supported by R35-HL135824 (Willer, PI). SMD is supported by IK2-CX001780. This research is based on data from the Million Veteran Program, Office of Research and Development, Veterans Health Administration, and was supported by award # BX003362. This publication does not represent the views of the Department of Veteran Affairs or the United States Government. This work was supported by funding from the Department of Veterans Affairs Office of Research and Development, Million Veteran Program Grant MVP000; Department of Veterans. The views expressed in this manuscript are those of the authors and do not necessarily represent the views of the National Heart, Lung, and Blood Institute; the National Institutes of Health; or the U.S. Department of Health and Human Services.

The Genotype-Tissue Expression (GTEx) Project was supported by the Common Fund of the Office of the Director of the National Institutes of Health, and by NCI, NHGRI, NHLBI, NIDA, NIMH, and NINDS. The data used for the analyses described in this manuscript were obtained from GTEx Analysis v8 on the GTEx Portal on 05/01/2021.

## Author Contributions

Bioinformatic analysis: WZ, BNW, IS, KHW

Mendelian randomization: BB, LB

Replication: FT, ADJ, NS, SD, DAT

Functional analysis: YZ, XY, CR, JS

Writing: BNW, IS, KHW, KD, CR, JS

Revision: CJW, MD, BN

## STAR Methods

### GWAS Meta-analysis

VTE is one of the pilot phenotypes of the Global Biobank Meta-analysis Initiative (GBMI)(Zhou et al., 2021a). VTE was defined as in the INVENT consortium(Klarin et al., 2017) and refined as in **Supplementary Table 8**. After phenotype harmonization, genome wide association study (GWAS) summary statistics across 9 international cohorts (BioMe, BioVU, CKB, ETBB, FinnGen, GNH, MGI, UCLA, UKB) with representation across five super populations (American [AMR], African [AFR], East Asian [EAS], Europeans [EUR, including Finnish and Non-Finnish European ancestries], South Asian [SAS]) were combined using inverse variance weighted meta-analysis with 27,987 cases, 1,035,290 controls, and an effective sample size of 107,409 (**Supplementary Table 9-11, Supplementary Figure 1**). We defined genome-wide significant loci as in (Zhou et al., 2021a) by iteratively spanning the ± 500 kb region around the most significant variant and merging overlapping regions until no genome-wide significant variants (p-value < 5e-8) were detected within ± 500 kb. The most significant variant in each locus was selected as the lead SNP. The genomic control factor (lambda) used to measure inflation of test statistics was 1.042 (**Supplementary Figure 7**) and therefore p-values were not adjusted for inflation.

### Replication

Replication was performed in a meta-analysis of International Network against Thrombosis (INVENT)(Lindström et al., 2019) and Million Veterans Program(Klarin et al., 2019) (MVP, version 4) including European, African and Hispanic ancestries (**Supplementary Table 12**). The meta-analysis was conducted using a fixed-effect model based on inverse-variance weighting, after variants with either low imputation quality (< 0.3) or low frequency (minor allele count < 5) were removed. Summary statistics were collected for the lead SNP at 38 genome-wide significant loci. In one case, the variant was not present, so the closest SNP with high linkage disequilibrium (LD) was used (r2>0.8): rs187506383 was used as a proxy for rs536995174. We considered the variant association a successful replication if the p-value was less than a Bonferroni corrected threshold of 0.0013. There are 1,994 cases and 15,148 controls part of eMERGE that were used in the INVENT meta-analysis and may overlap with the BioVU cohort used in the GBMI meta-analysis. As a sensitivity analysis, GBMI summary statistics without BioVU were also compared to the replication summary statistics.

### Statistical analysis

Unless otherwise noted, analysis was performed in R. Packages for data visualization and statistical tests included: isoplotR(Vermeesch, 2018) ggplot2(Wickham, 2016), MASS(Venables WNRipley), data.table(Dowle and Srinivasan, 2021), ggthemes(Arnold, 2016), dplyr(Wickham et al., 2022). Code is available https://github.com/bnwolford/gbmi_vte.

### Credible sets

To fine-map the loci identified in the VTE meta-analysis, we generated credible sets of causal variants with SUm of SIngle Effects (SuSiE)(Wang et al., 2020) an iterative Bayesian step-wise selection using sparse multiple regression-determined credible sets with 95% posterior probability of containing potential causal variants. An LD reference panel from 2,504 unique individuals from all ancestral cohorts of 1000 Genomes was used. Regions containing likely causal variants to create credible sets were defined by +/- 500 kb from the index variant.

### Gene prioritization

For each autosomal locus we performed gene prioritization using seven independent, yet converging, lines of evidence. We performed DEPICT and PoPS for gene prioritization for all 14 endpoints in the GBMI pilot study(Zhou et al., 2021a). Using the variants with p-value <1e-5 in the multi-ancestry meta-analysis, any gene with p-value <0.05 FDR with DEPICT(1000 Genomes Project Consortium et al., 2010; Pers et al., 2015) was considered prioritized. Similarly, a gene in the top 10% of genes ranked by PoPS(Weeks et al., 2020) was considered as the prioritized genes. For both analyses, individuals of European ancestry from 1000 Genomes Project phase 3 were used as the LD reference panel(1000 Genomes Project Consortium et al., 2015), since 86% of the individuals included in the GWAS were of primarily-European ancestry.

We compared the performance of DEPICT and PoPS using a ‘gold standard’ set of VTE genes (N=41, **Supplementary Table 2**), determined prior to the GWAS by a medical and molecular genetics expert in VTE and coagulation (J. Shavit) and based on a high throughput sequencing panel by the ThromboGenomics group(Downes et al., 2019). We used a significant threshold of FDR<0.05 to define prioritized genes from the DEPICT result. Of the 54 genes prioritized by DEPICT, 11 of those were in the VTE gold standard gene list (AUC=0.75, **Supplementary Figure 8**). For the PoPS gene prioritization result, we selected the top 10% genes (n=1,839) with the highest PoP score as the prioritized genes, and 32 of the PoPS defined prioritized genes were reported in the gold standard gene list (AUC=0.84, **Supplementary Figure 8**). We further evaluated the performance between DEPICT and PoPS in predicting functional genes with the DeLong test, which showed no significant difference between the accuracy of the two methods (p-value=0.30). Therefore we concluded both methods in the integrative prioritization.

For the Proteome-wide Mendelian randomization (PWMR) (Zhao et al. 2022) and colocalization analysis, candidate SNPs from the NFE meta-analysis were selected with p-value < 1×10^−5^ for genetic association with VTE. For approximate colocalization, we aimed to test whether the leading pQTL was in LD (r^2^ ≥ 0.8) `with a candidate SNP. Genes with PWMR and co-localization evidence were used for` prioritization. We also looked up the lead SNPs in relevant GTEx tissues(GTEx Consortium, 2020)—whole blood, atrial appendage, left ventricle, aortic artery, coronary artery, tibial artery, EBV-transformed lymphocytes— and reported significant eQTLs (q-value < 0.05).

We also considered deleterious mutations for gene prioritization. We identified genes with a pathogenic variant in ClinVar(Landrum et al., 2018) as of 05-07-2021 or genes with a nonsynonymous variant in the 95% credible set (calculated with sum of single effects SuSiE). Finally, we considered the nearest gene as annotated with ANNOVAR. For each lead SNP, a simple sum across these seven lines of evidence was used to identify a potentially causal gene with the most evidence. We used Enrichr(Chen et al., 2013; Kuleshov et al., 2016; Xie et al., 2021) for the 38 prioritized candidate genes (one per locus) to identify Gene Ontology (GO) Molecular Function and Biological Processes and KEGG Human pathways.

### Mendelian Randomization

We performed two-sample Mendelian randomization to investigate the causal relation of VTE with COVID hospitalization. The NFE ancestry specific summary statistics for VTE were used with an effective sample size of 61678 (cases=15,970, controls=681,106). The European ancestry specific summary statistics of COVID-19 hospitalization were accessed from the COVID-19 Host Genetics Initiative(COVID-19 Host Genetics Initiative, 2021) with a sample size 1,835,054 (cases=18,061, controls=1,816,993). LD-clumping of summary statistics of VTE was performed using PLINK v1.9 (Chang et al., 2015; [CSL STYLE ERROR: reference with no printed form.]) to identify the independent 20 significant COVID-19-associated SNPs using thresholds p-value<5×10^−8^, r^2^=0.001, and kb=10000. After matching and harmonization of 20 VTE SNPs effect with COVID-19 summary statistics, rs10142521 and rs1799810 were removed for being palindromic with intermediate allele frequencies. We used 18 SNPs as an instrument for VTE (**Supplementary Table 13**) and applied inverse-variance weighted, MR-Egger, weighted median, penalized weighted median, weighted mode, and pleiotropy residual sum and outlier methods using MR-Base and MR-PRESSO(Verbanck et al., 2018). In the MR-PRESSO analysis, the three SNPs rs10901263, rs2289252, and rs78707713 were excluded as they were detected as SNP effect size outliers in the main analysis. For African (AFR) and Hispanic (HIS) specific analysis, we did not have enough independent significant SNPs (AFR=2, HIS=0) for VTE to perform analyses in multiple ancestries. All the analyses were performed using TwoSampleMR(Hemani et al., 2017, 2018) package in R.

### Targeted mutagenesis in zebrafish using genome engineering

We used the ChopChop server to identify highly efficient single guide (RNAs) sgRNAs for CRISPR/Cas9 mediated genome editing(Labun et al., 2019). Two to four guides were selected for each gene of interest (**Supplementary Table 14**). sgRNAs and Cas9 nuclease were ordered from Synthego. 1.5 nanomoles (nmol) of each sgRNA from Synthego were mixed with 15 microliters (ul) of nuclease-free water, and then immediately aliquoted and stored at -80°C. During each injection, 2 ul of the original aliquoted sgRNA was diluted with 31 ul of nuclease-free water. Cas9 solution was prepared by combining 5.37 ul of water, 1.54 ul of Cas9 nuclease, 1.78 ul of 0.1M HEPES, and 1.33 ul of 1M KCL. The diluted sgRNAs were mixed with the Cas9 solution in a 1:1 volume ratio. This mix was incubated at 37°C for 10 minutes, and 4 to 6 nL were injected into the yolk of single-cell embryos produced from ABxTL F1 hybrid adults. Uninjected sibling clutchmates were set aside as controls. Solution without sgRNA was also injected as a negative control.

### Laser-induced endothelial injury in zebrafish

Zebrafish were maintained according to protocols approved by the University of Michigan Animal Care and Use Committee. Three days post fertilization (dpf) larvae of both injected and uninjected clutchmate controls were anesthetized with tricaine and mounted in 1.6% low melting agarose. Laser injury was performed under an Olympus IX73 inverted microscope using an Andor Micropoint pulsed-dye laser and focusing system. 99 pulses were administered to the posterior cardinal vein (PCV), 5 somites caudal to the anal pore(Liu et al., 2014; Rost et al., 2016). Following injury, larvae were observed until complete occlusion of the vessel and the time to occlusion (TTO) was recorded. Larvae that did not occlude within 2 minutes were assigned a TTO of 120 seconds. After laser injury, larvae were anesthetized and individually processed with lysis buffer (10 mM Tris-Cl, pH 8.0; 2 mM EDTA, 2% Triton X-100, and 100 µg/mL Proteinase K) at 55°C for at least 30 minutes, followed by heat inactivation at 95°C for 5 minutes. Crude lysate was subjected to polymerase chain reaction (PCR) using primers flanking each sgRNA target site to confirm successful genome editing.

Statistical analysis was performed with the ggpubr package in R v4.1.1. Pairwise Wilcoxon signed rank test was performed for each gene to compare time to occlusion of uninjected controls and sgRNA injected embryos. Injection with no sgRNA was used as a negative control. A sensitivity analysis was performed by pooling uninjected control measurements and comparing this distribution to that of each sgRNA injected gene using Wilcoxon signed rank test. A Bonferroni threshold of 0.0083 was used to account for six independent tests.

