## Supplementary material for "Multi-ancestry GWAS for venous thromboembolism identifies novel loci followed by experimental validation in zebrafish": GBMI-VTE-Supplemental-Material

### Supplementary Information

**Supplementary Figure 1. Sample sizes across studies and ancestries**
The left x-axis is showing VTE (venous thromboembolism) prevalence across the biobanks shaded by sampling strategy and the right x-axis is showing the number of VTE cases across the biobanks shaded by global ancestry labels.
UCLA=UCLA Precision Health Biobank; MGI=Michigan Genomics Initiative; BioMe=Mount Sinai BioMe Biobank; BioVU=Biorepository at Vanderbilt University; UKBB=United Kingdom Biobank; ESTBB=Estonia Biobank; GNH=East London Genes and Health; CKB=China Kadoorie Biobank; AFR= African ancestry; MID=Middle Eastern ancestry; AMR=Admixed American ancestry; EAS=East Asian ancestry; NFE=non-Finnish European; FIN=Finnish ancestry; SAS=South Asian ancestry

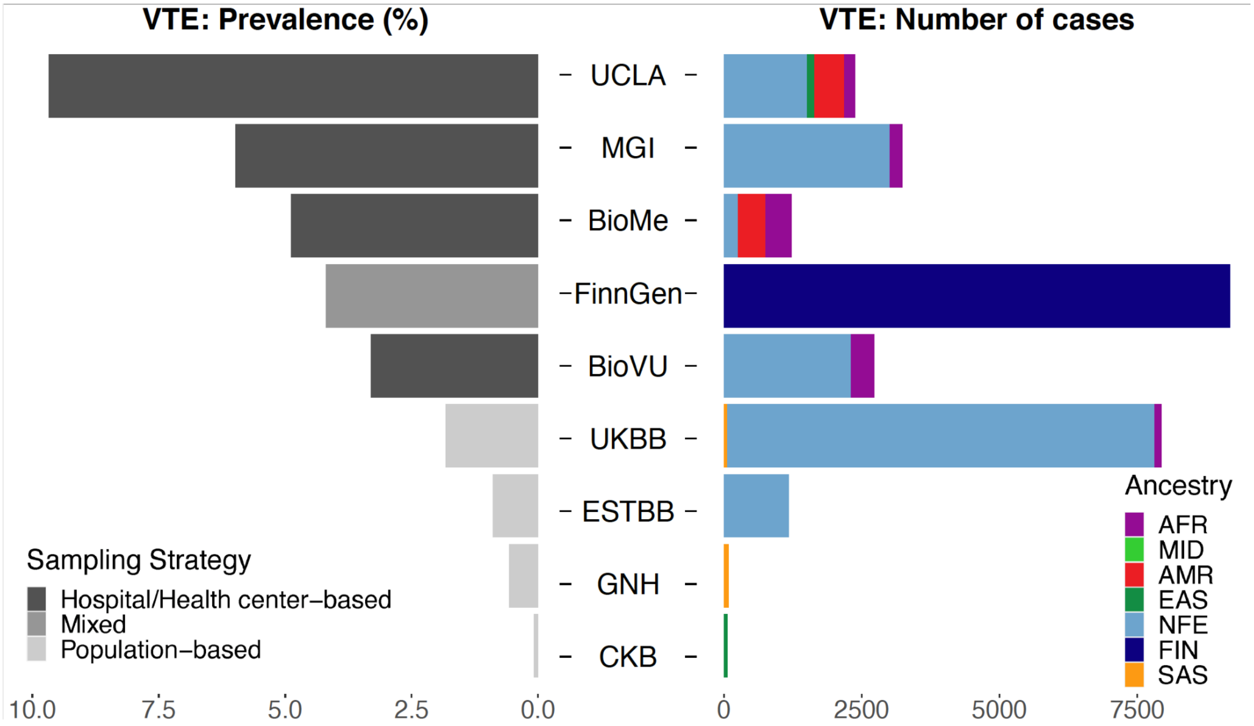

**Supplementary Figure 2. Differential effect sizes between population-based and hospital-based biobank meta-analyses**

Meta-analysis in only population-based biobanks (UK Biobank, Estonian Biobank, Genes and Health, China Kadoorie Biobank) and hospital-based biobanks (UCLA Precision Health Biobank, Michigan Genomics Initiative, Mount Sinai BioMe Biobank, Biorepository at Vanderbilt University ) was performed for index variants of genome wide significant loci for VTE. Only 4 loci have a p-value below $1x10^{-6}$ in both population-based and hospital-based meta-analyses. Overall, effect sizes are larger for population-based biobanks than hospital-based biobanks (slope=1.4, 95% CI 1.2-1.6).

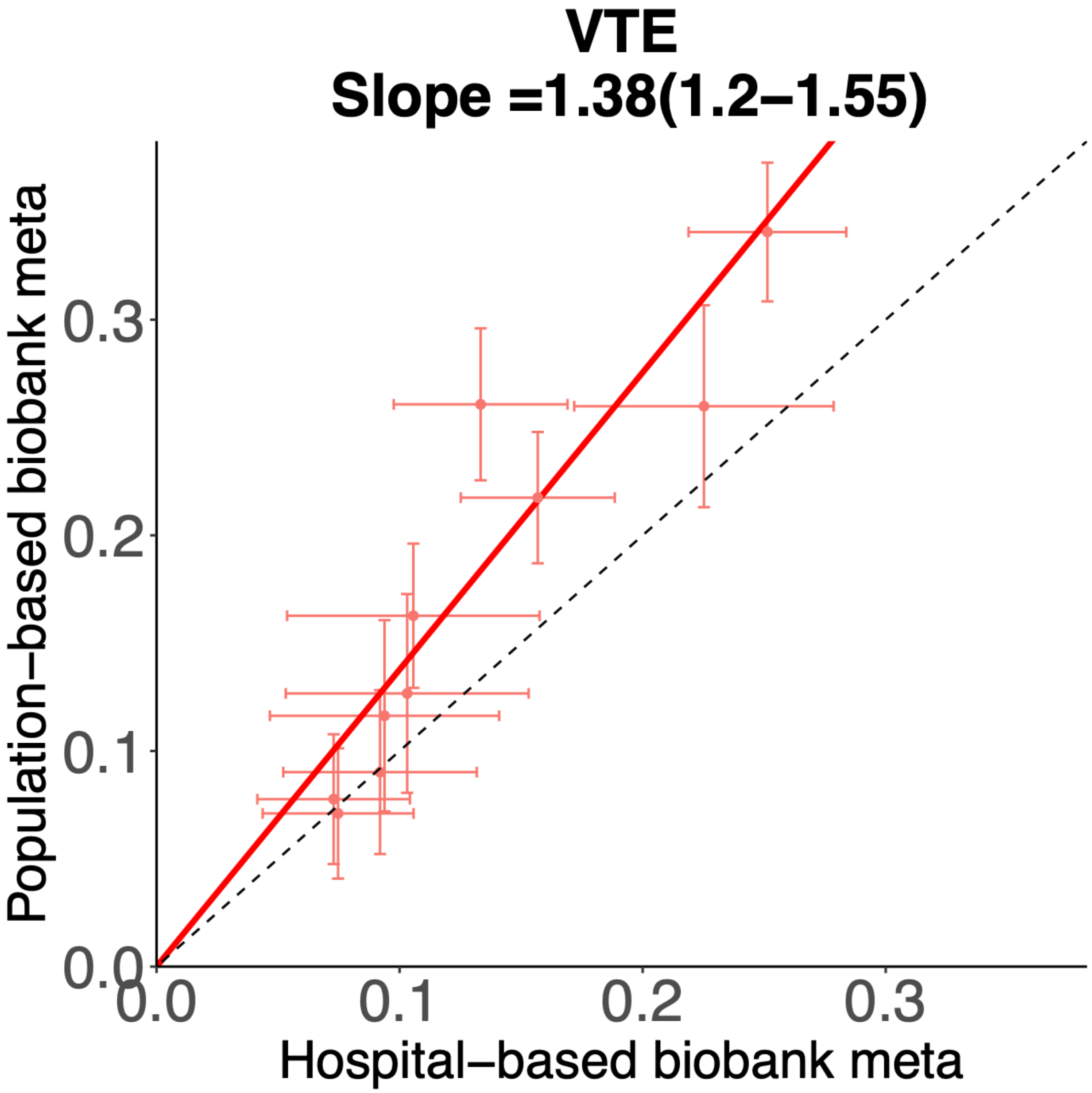

**Supplementary Figure 3. Variants with nominally significant heterogeneity across ancestries**

Three variants (x-axis) had nominally (p-value < 0.05) significant heterogeneity across contributing ancestries. The estimated effect sizes (y-axis) shown here grouped by ancestry (NFE=non Finnish European; FIN=Finnish; AFR=African; ALL=combined ancestries in meta-analysis)

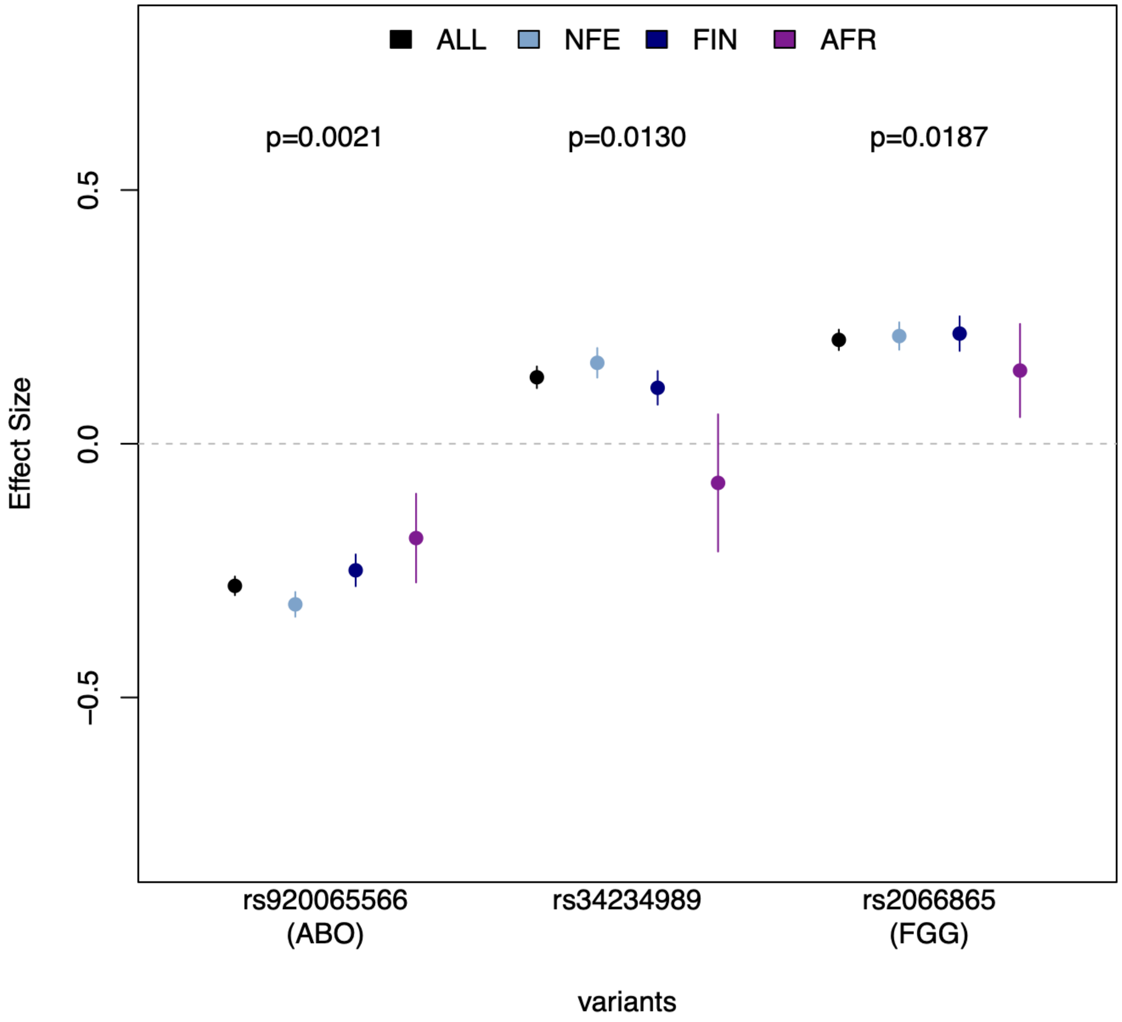

**Supplementary Figure 4. Ancestry specific locus rs112106699 near *DHRS3***

Summary statistics and forest plot for contributing cohorts and meta-analysis (in bold) at one genome wide significant variant. PoPS prioritized gene *DHRS3* is 4.4kilobases (kb) from the tag SNP and *VPS13D* is 51.4 kb away.

Freq=effect alelle frequency; Beta=estimated effect size from GWAS; BioMe_afr=Mount Sinai BioMe Biobank African ancestry; BioVU_afr=Biorepository at Vanderbilt University African ancestry; UKBB_afr=UK Biobank African ancestry; BioME_amr=Mount Sinai BioMe Biobank American ancestry; MGI_nfe=Michigan Genomics Initiative non-Finnish European ancestry; UKBB_nfe=UK Biobank non-Finnish European; UKBB_sas=UK Biobank South Asian ancestry

| Biobank_Ancestry | Freq | Beta | P−value |
|---|---|---|---|
| BioMe_afr | 0.0906 | 0.5228 | 5.66e−03 |
| BioVU_afr | 0.0599 | 0.3738 | 3.75e−03 |
| MGI_afr | 0.0806 | 0.4379 | 2.62e−02 |
| UKBB_afr | 0.0986 | 0.4687 | 3.29e−02 |
| BioMe_amr | 0.0258 | 0.4987 | 1.14e−01 |
| MGI_nfe | 0.0014 | 0.6274 | 2.59e−01 |
| UKBB_nfe | 5e−04 | 0.8431 | 7.03e−02 |
| UKBB_sas | 0.0031 | 1.4447 | 5.31e−01 |
| **Summary** | **0.0045** | **0.4538** | **2.04e−08** |

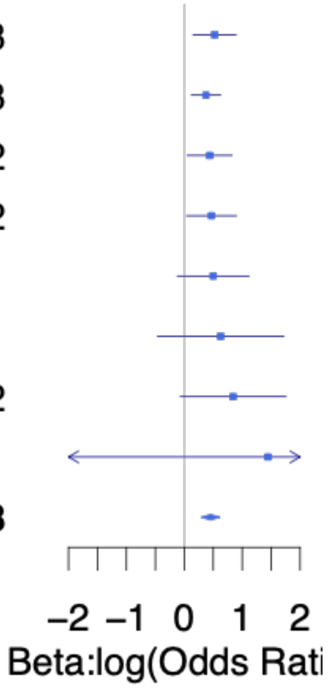

**Supplementary Figure 5. Scatterplot of effect sizes between discovery and replication datasets**

**A.**

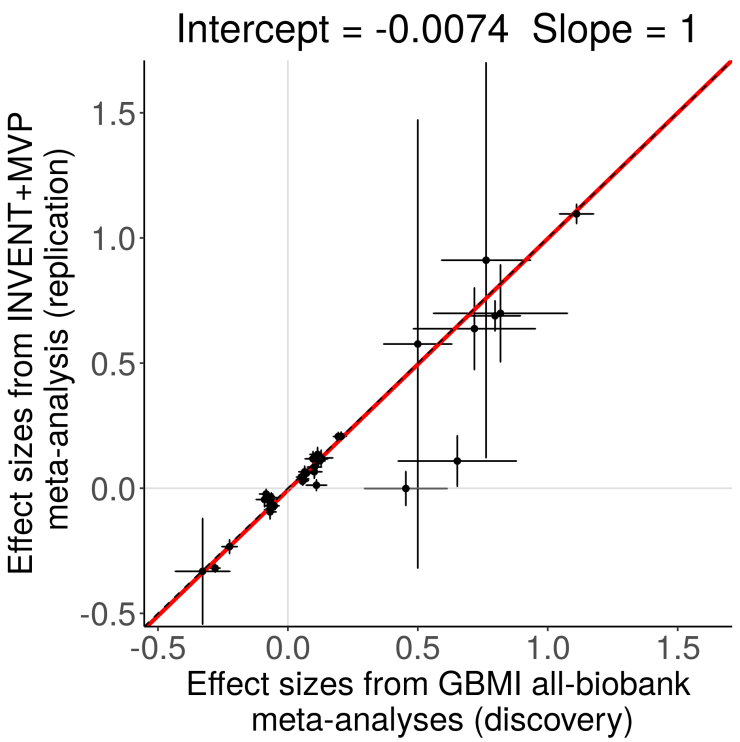


**B.**

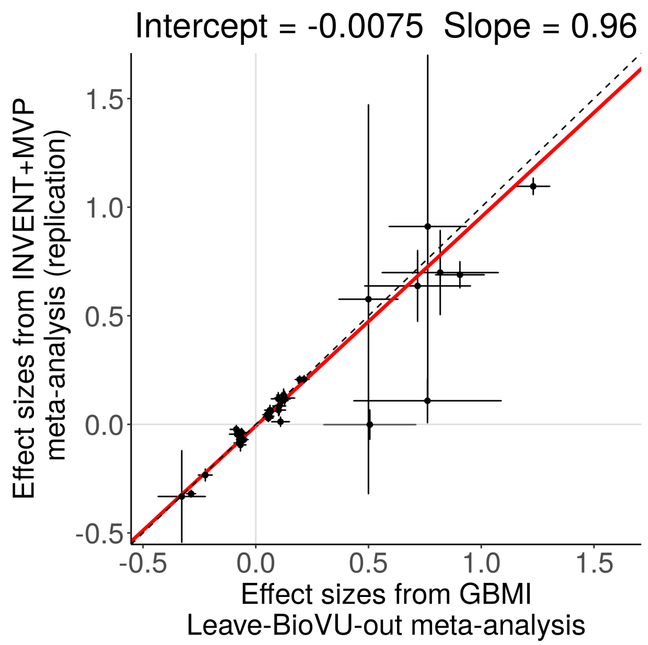

a. Scatterplot of effect sizes with standard error bars for INVENT+MVP (y-axis) versus GBMI meta-analysis (x-axis). Intercept and slope shown are from York's regression (see STAR methods).
b. Scatterplot of effect sizes with standard error bars for INVENT+MVP (x-axis) versus GBMI meta-analysis without BioVU. This is a sensitivity analysis since the INVENT meta-analysis included eMERGE samples that could overlap with those of BioVU.

**Supplementary Figure 6. Functional experiment pooling non-injected controls**
Boxplot of time to occlusion in seconds (y-axis) for pooled uninjected controls (purple) and targeted genes (injected). P-values from wilcoxon rank sum test comparing all uninjected controls to each targeted gene are shown for each paired test.
sgRNA=single guide RNA

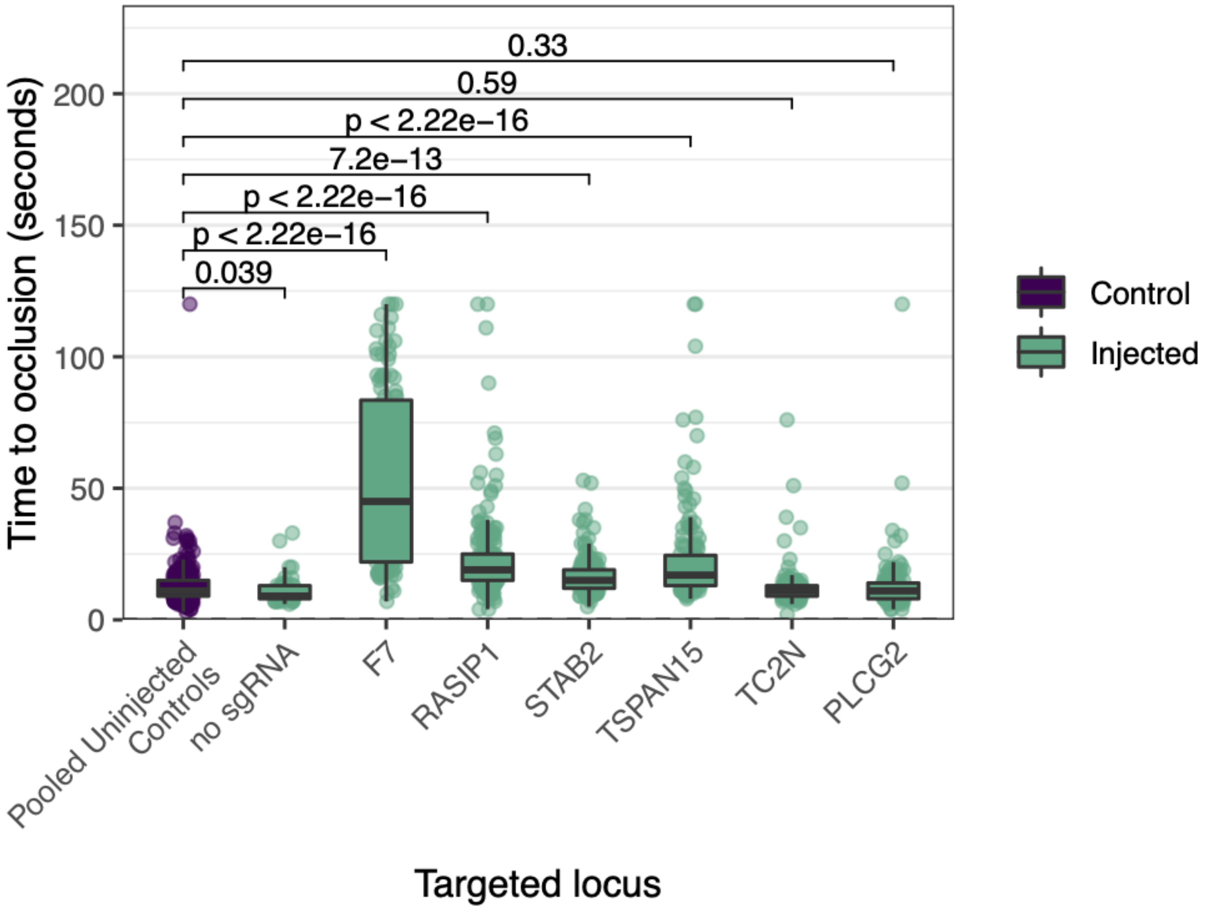

**Supplementary Figure 7. Quantile-quantile plot (QQplot) for meta-analysis**

Quantile-Quantile plot (QQplot) for meta-analysis p-values grouped and colored by minor allele frequency (MAF). Lambda at quantiles (0.001, 0.01, 0.1, 0.5, and 0.7) are calculated.

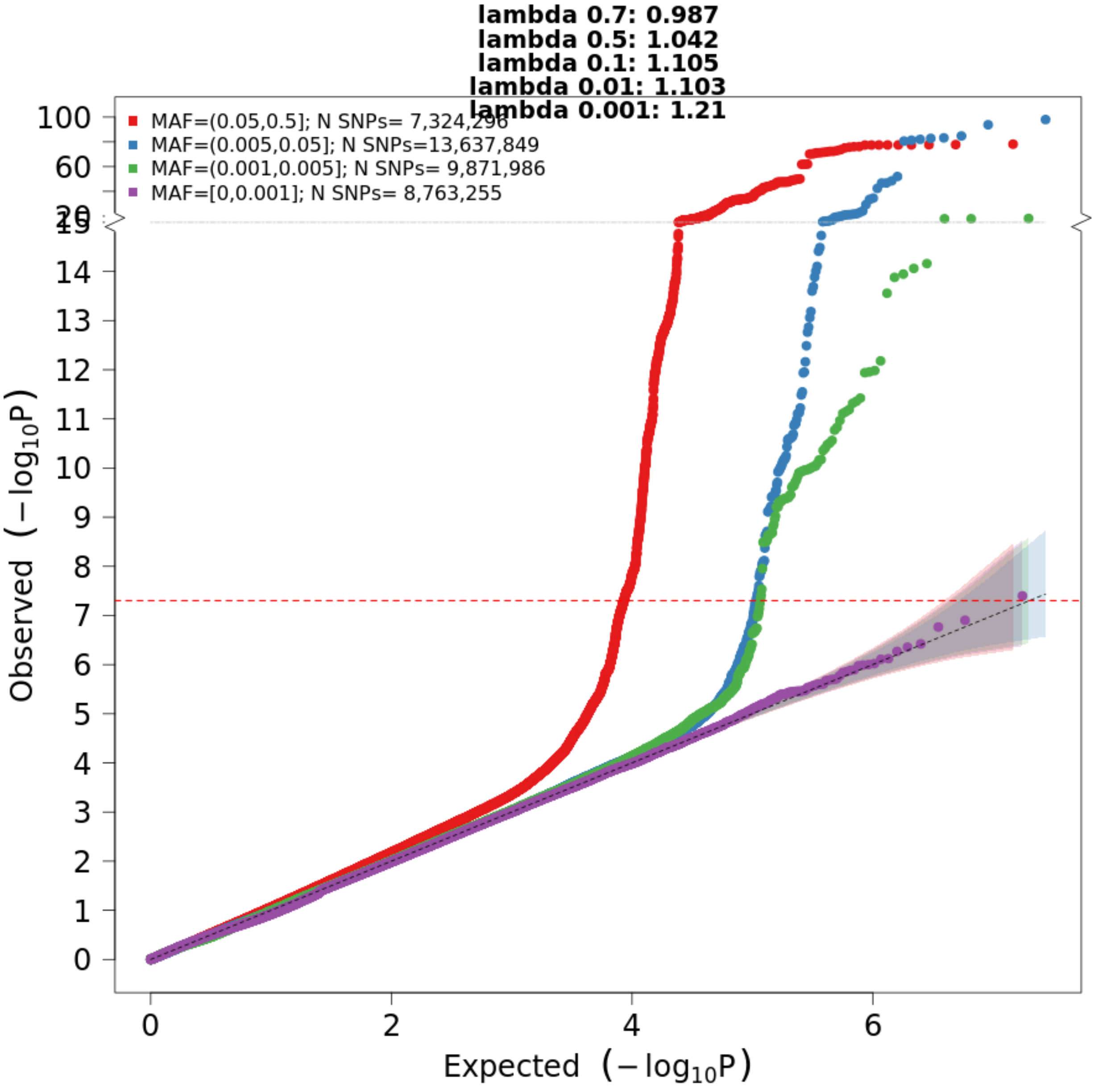

**Supplementary Figure 8. Comparison of DEPICT and PoPs using a gold standard gene set**
Specificity (x-axis) and sensitivity (y-axis) for prioritization of 41 gold standard VTE genes using DEPICT (reds) and PoPS (blues). The multi-ancestry meta-analysis was used (ALL) as well as the non-Finnish European ancestry meta-analysis (NFE).

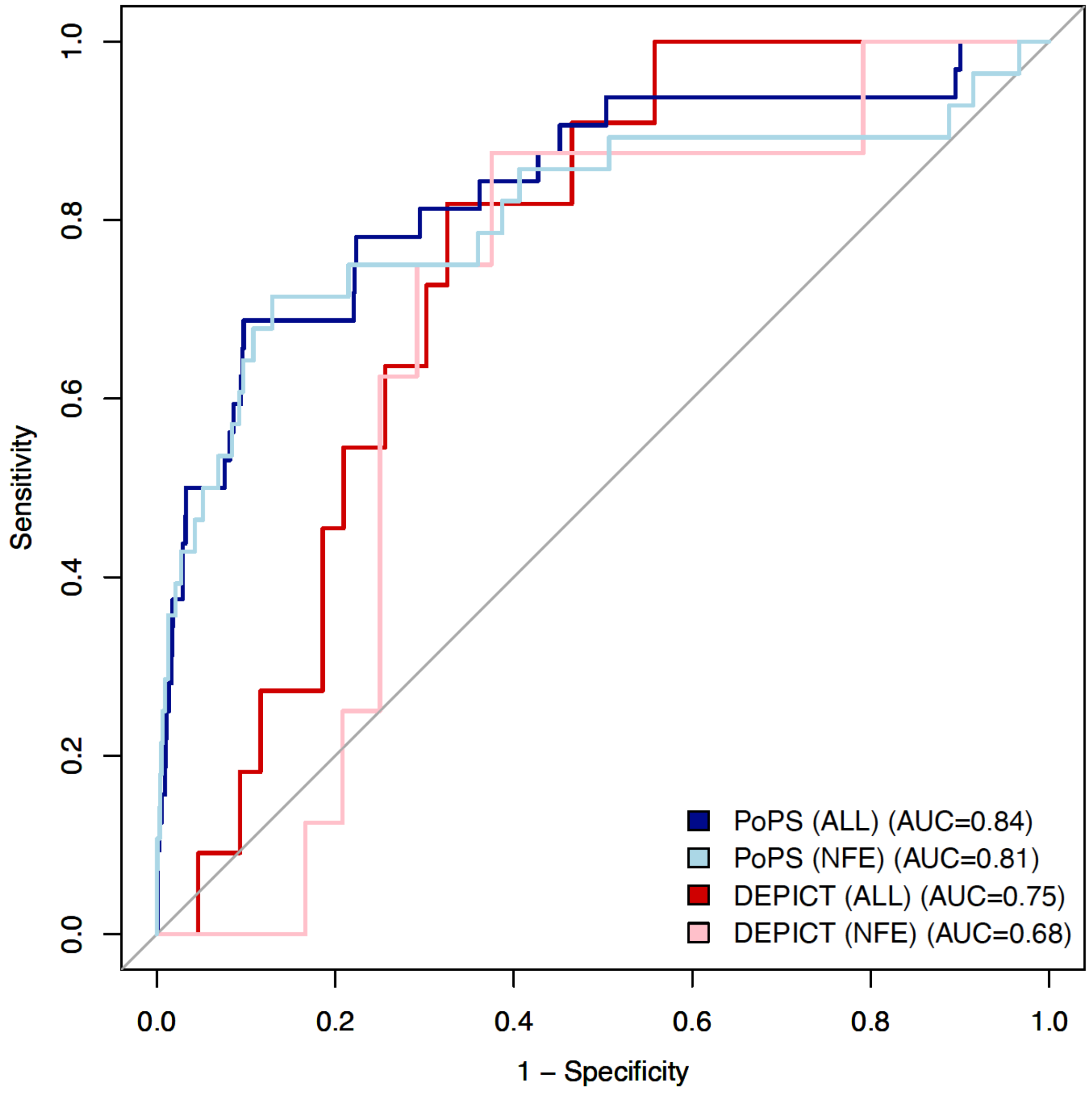

**Supplementary Table 1: Previous VTE studies from the NHGRI-EBI GWAS catalog as of October 2021**

Accompanying excel file

* GWAS for platelet thrombin activation

** GWAS for factor 5 plasma levels

Genome-wide association studies for Venous Thromboembolism in the GWAS Catalog (Buniello et al., 2019) as of June 2022. References: (Trégouët et al., 2009) (Germain et al., 2011) (Heit et al., 2012) (Greliche et al., 2013) (Tang et al., 2013) (Germain et al., 2015) (Hernandez et al., 2016) (Hinds et al., 2016) (Rühle et al., 2017) (Heit et al., 2017) (Klarin et al., 2017) (Thibord et al., 2019) (Klarin et al., 2019) (Deguchi et al., 2020) (Rodriguez et al., 2020) (Mateos et al., 2020) (Herrera-Rivero et al., 2021)

**Supplementary Table 2. Gold standard coagulation genes with associations with Venous Thromboembolism**

Genes known to be associated with blood coagulation and/or platelet disorder functional assays and human genetics assays known from the literature(Downes et al., 2019). The list is filtered for genes that are not duplicated in zebrafish for experimental purposes. Notably, the landscape of gold standard genes is slightly different from the genes identified via genome wide association studies (GWASs).

Accompanying excel file

* Also associated with platelet disorder

**Supplementary Table 3. 38 genome-wide significant loci from GBMI meta-anlaysis**

Accompanying excel file

31 lead SNPs are within 500 kb of a variant previously reported in GWAS or sequencing studies, resulting in 9 potentially novel associations

**Supplementary Table 4. Meta-analysis of replication and leave BioVU out GBMI meta-analysis**

Accompanying excel file

Of the 37 markers available for replication in the INVENT+MVP meta-analysis, 34 are genome-wide significant ($5x10^{-8}$) with inverse variance based meta-analysis and two sided p-value. This sensitivity analysis accounts for possible inclusion of samples in both the INVENT+MVP analysis and the GBMI meta-analysis with BioVU included.

**Supplementary Table 5. KEGG Pathways**

Accompanying excel file

Kyoto Encyclopedia of Genes and Genomes (KEGG, Human cell signaling and metabolic pathways version 2021) enriched in 38 prioritized genes from genome-wide significant loci. Adjusted p-value is using Benjamini-Hochberg method for multiple hypothesis testing.

**Supplementary Table 6. Gene Ontology (GO) Biological Processes**

Accompanying excel file

Gene Ontology (GO) terms for biological processes (2021 version) enriched in 38 prioritized genes from genome-wide significant loci. Adjusted p-value is using Benjamini-Hochberg method for multiple hypothesis testing.

**Supplementary Table 7. Gene Ontology (GO) Molecular Function**

Accompanying excel file

Gene Ontology (GO) terms for biological processes (2018 version) enriched in 38 prioritized genes from genome-wide significant loci. Adjusted p-value is using Benjamini-Hochberg method for multiple hypothesis testing.

**Supplementary Table 8: Case and control definitions used in analysis plan provided to participating biobanks**

Note, the replication cohort INVENT defined also included cases from self-report which were interview with a trained nurse to confirm diagnosis(Klarin et al., 2017).

| Subset | ICD codes | Description |
|---|---|---|
| cases inclusion (ICD10) | I80.1, I80.2, I82.2 , I26.0, I26.9 | Hospitalization for ICD-10 Code I80.1 or I80.2 - phlebitis and thrombophlebitis of the femoral vein or other deep vessels of lower extremities;<br><br>Hospitalization for ICD-10 Code I82.2 - embolism and thrombosis of vena cava;<br><br>Hospitalization for ICD-10 Code I26.0 or I26.9 - pulmonary embolism with or without acute cor pulmonale; |
| cases inclusion (ICD9) | 451.11, 453.40, 453.2, 453.77, 453.87, 415.1 | |
| cases inclusion (OPCS codes) | L79.1 or L90.2 | Hospitalization for OPCS-4 Procedures Codes L79.1 or L90.2 - open venous thrombectomy of lower limb vein or inferior vena cava filter insertion.<br>Open venous thrombectomy of lower limb vein or inferior vena cava filter insertion. |
| controls exclusion (ICD10) | I81, I82.0, I80.0, I80.3, I80.8, I80.9, D68 | portal vein thrombosis (I81), Budd-Chiari syndrome (I82.0), superficial or unclear site of thrombophlebitis (I80.0, I80.3, I80.8, I80.9), or known coagulation defects (D68). |
| controls exclusion (ICD9) | 452,453.0,451.0,451.89,451.9, 286 | |

**Supplementary Table 9: Effective sample sizes across ancestries**

UCLA=UCLA Precision Health Biobank; MGI=Michigan Genomics Initiative; BioMe=Mount Sinai BioMe Biobank; BioVU=Biorepository at Vanderbilt University; UKBB=United Kingdom Biobank; ESTBB=Estonia Biobank; GNH=East London Genes and Health; CKB=China Kadoorie Biobank

| **Ancestries** | **Cohorts** | **Cases** | **Controls** | **Effective Sample Size** |
|---|---|---|---|---|
| African | BioMe, BioVU, UKB, UCLA | 1,466 | 31,042 | 5,477 |
| Admixed American | BioMe, UCLA | 1,037 | 12,479 | 3,772 |
| East Asian | CKB, UCLA | 193 | 77,462 | 741 |
| Finnish | FinnGen | 9,176 | 209,616 | NA |
| Non-Finnish European | BioMe, BioVU, EstBB, MGI, UCLA, UKB | 15,970 | 681,106 | 61,678 |
| South Asian | GNH, UKB | 145 | 23,585 | 576 |
| **Total** | | **27,987** | **1,035,290** | **107,409** |

**Supplementary Table 10. Sample size for VTE in each cohort by ancestry, sexes combined**
UCLA=UCLA Precision Health Biobank; MGI=Michigan Genomics Initiative; BioMe=Mount Sinai BioMe Biobank; BioVU=Biorepository at Vanderbilt University; UKBB=United Kingdom Biobank; ESTBB=Estonia Biobank; GNH=East London Genes and Health; CKB=China Kadoorie Biobank; AFR= African ancestry; MID=Middle Eastern ancestry; AMR=Admixed American ancestry; EAS=East Asian ancestry; NFE=non-Finnish European; FIN=Finnish ancestry; SAS=South Asian ancestry

| **Biobank** | **Ancestry** | **Case** | **Control** |
|---|---|---|---|
| BioMe | afr | 477 | 6287 |
| BioMe | nfe | 253 | 8903 |
| BioMe | amr | 494 | 8647 |
| BioVU | afr | 434 | 14295 |
| BioVU | nfe | 2295 | 65621 |
| CKB | eas | 61 | 75294 |
| ESTBB | nfe | 1177 | 131802 |
| FinnGen | fin | 9176 | 209616 |
| GNH | sas | 84 | 14793 |
| MGI | afr | 231 | 2935 |
| MGI | nfe | 2999 | 47845 |
| UCLA | afr | 195 | 1031 |
| UCLA | amr | 543 | 3832 |
| UCLA | eas | 132 | 2168 |
| UCLA | nfe | 1503 | 15138 |
| UKBB | afr | 129 | 6494 |
| UKBB | sas | 61 | 8792 |
| UKBB | nfe | 7743 | 411797 |
| Total | | 27987 | 1035290 |

**Supplementary Table 11. Effective Sample Sizes for each cohort, by ancestries, by sex**
Accompanying excel file
Effect sample sizes for each cohort, each ancestry group for both sexes combined and stratified by sex. Sample names with "leave" are for each leave one biobank out meta-analysis performed. UCLA=UCLA Precision Health Biobank; MGI=Michigan Genomics Initiative; BioMe=Mount Sinai BioMe Biobank; BioVU=Biorepository at Vanderbilt University; UKBB=United Kingdom Biobank; ESTBB=Estonia Biobank; GNH=East London Genes and Health; CKB=China Kadoorie Biobank; AFR= African ancestry; MID=Middle Eastern ancestry; AMR=Admixed American ancestry; EAS=East Asian ancestry; NFE=non-Finnish European; FIN=Finnish ancestry; SAS=South Asian ancestry

**Supplementary Table 12. Replication cohort sample sizes by ancestry groups**
INVENT=International Network on Venous Thrombosis
MVP=Million Veteran Program

| **Cohort and ancestry** | **Cases** | **Controls** |
|---|---|---|
| INVENT European | 15,572 | 113,430 |
| INVENT African | 799 | 14,353 |
| MVP European | 18,952 | 411,746 |
| MVP African | 5,521 | 103,791 |
| MVP Hispanic | 1,188 | 45,634 |
| **Total** | 42,032 | 688,954 |

**Supplementary Table 13. Genetic instruments for two-sample Mendelian Randomization**
Accompanying excel file
18 single nucleotide polymorphisms (SNPs) selected as genetic instruments.
EAF=effect allele frequency; SE=standard error

**Supplementary Table 14. Efficient single guide RNAs (sgRNA) for CRISPR/Cas9 mediated genome editing.**
Accompanying excel file
Genes of interest identified for validation in zebrafish and the corresponding sgRNAs used for CRISPR/Cas9 genome editing knockdown.

**GBMI Membership**

Wei Zhou[1,2,3], Masahiro Kanai[1,2,3,4,5], Kuan-Han H Wu[6], Humaira Rasheed[7,8,9], Kristin Tsuo[1,2,3], Jibril B Hirbo[10,11], Ying Wang[1,2,3], Arjun Bhattacharya[12], Huiling Zhao[9], Shinichi Namba[5], Ida Surakka[13], Brooke N Wolford[6,7], Valeria Lo Faro[14,15,16], Esteban A Lopera- Maya[17], Kristi Läll[18], Marie-Julie Favé[19], Sinéad B Chapman[2,3], Juha Karjalainen[1,2,3,20], Mitja Kurki[1,2,3,20], Maasha Mutaamba[1,2,3,20], Juulia Partanen[20], Ben M Brumpton[7,21,22], Sameer Chavan[23], Tzu-Ting Chen[24], Michelle Daya[23], Yi Ding[12,25], Yen-Chen A Feng[26,27], Christopher R Gignoux[23], Sarah E Graham[13], Whitney E Hornsby[13], Nathan Ingold[28,29], Ruth Johnson[12,30], Triin Laisk[18], Kuang Lin[31], Jun Lv[32], Iona Y Millwood[31,33], Priit Palta[18,20], Anita Pandit[34], Michael H Preuss[35], Unnur Thorsteinsdottir[36], Jasmina Uzunovic[19], Matthew Zawistowski[34], Xue Zhong[10,11], Archie Campbell[37], Kristy Crooks[23], Geertruida H de Bock[38], Nicholas J Douville[39,40], Sarah Finer[41], Lars G Fritsche[34], Christopher J Griffiths[41], Yu Guo[42], Karen A Hunt[43], Takahiro Konuma[5,44], Riccardo E Marioni[37], Jansonius Nomdo[14], Snehal Patil[34], Nicholas Rafaels[23], Anne Richmond[45], Jonathan A Shortt[23], Peter Straub[10,11], Ran Tao[11], Brett Vanderwerff[34], Kathleen C Barnes[23], Marike Boezen‡[38], Zhengming Chen[31,33], Chia-Yen Chen[46], Judy Cho[35], George Davey Smith[9,47], Hilary K Finucane[1,2,3], Lude Franke[17], Eric R Gamazon[10,11,48], Andrea Ganna[1,2,20], Tom R Gaunt[9], Tian Ge[27,49], Hailiang Huang[1,2], Jennifer Huffman[50], Jukka T. Koskela[20], Clara Lajonchere[51,52], Matthew H Law[28,29], Liming Li[32], Cecilia M Lindgren[53], Ruth JF Loos[35,54], Stuart MacGregor[28], Koichi Matsuda[55], Catherine M Olsen[28], David J Porteous[37], Jordan A Shavit[56], Harold Snieder[38], Richard C Trembath[57], Judith M Vonk[38], David Whiteman[28], Stephen J Wicks[23], Cisca Wijmenga[17], John Wright[58], Jie Zheng[9], Xiang Zhou[34], Philip Awadalla[19,59], Michael Boehnke[34], Nancy J Cox[10,11], Daniel H Geschwind[51,60,61], Caroline Hayward[45], Kristian Hveem[7,21], Eimear E Kenny[62], Yen-Feng Lin[24,63,64], Reedik Mägi[18], Hilary C Martin[65], Sarah E Medland[28], Yukinori Okada[5,66,67,68,69], Aarno V Palotie[1,2,20], Bogdan Pasaniuc[12,25,51,60,70], Serena Sanna[17,71], Jordan W Smoller[27], Kari Stefansson[36], David A van Heel[43], Robin G Walters[31,33], Sebastian Zöllner[34], Biobank Japan, BioMe, BioVU, Canadian Partnership for Tomorrow's Health/Ontario Health Study, China Kadoorie Biobank Collaborative Group, Colorado Center for Personalized Medicine, deCODE Genetics, Estonian Biobank, FinnGen, Generation Scotland, Genes & Health, LifeLines, Mass General Brigham Biobank, Michigan Genomics Initiative, QIMR Berghofer Biobank, Taiwan Biobank, The HUNT Study, UCLA ATLAS Community Health Initiative, UK Biobank, Alicia R Martin[1,2,3], Cristen J Willer[6,13,72]*, Mark J Daly[1,2,3,20]*, Benjamin M Neale[1,2,3]*

‡Deceased
*These authors jointly supervised this work

[1]Analytic and Translational Genetics Unit, Department of Medicine, Massachusetts General Hospital, Boston, MA, USA, [2]Stanley Center for Psychiatric Research, Broad Institute of MIT and Harvard, Cambridge, MA, USA, [3]Program in Medical and Population Genetics, Broad Institute of MIT and Harvard, Cambridge, MA, USA, [4]Department of Biomedical Informatics, Harvard Medical School, Boston, MA, USA, [5]Department of Statistical Genetics, Osaka University Graduate School of Medicine, Suita 565-0871, Japan, [6]Department of Computational Medicine and Bioinformatics, University of Michigan, Ann Arbor, MI, USA, [7]K.G. Jebsen Center for Genetic Epidemiology, Department of Public Health and Nursing, NTNU, Norwegian University of Science and Technology, Trondheim, Norway, [8]Division of Medicine and

Laboratory Sciences, University of Oslo, Norway, [9]MRC Integrative Epidemiology Unit (IEU), Bristol Medical School, University of Bristol, Bristol, UK, [10]Department of Medicine, Division of Genetic Medicine, Vanderbilt University Medical Center, Nashville, TN, USA, [11]Vanderbilt Genetics Institute, Vanderbilt University Medical Center, Nashville, TN, USA, [12]Department of Pathology and Laboratory Medicine, David Geffen School of Medicine, University of California, Los Angeles, Los Angeles, CA, USA, [13]Department of Internal Medicine, Division of Cardiology, University of Michigan, Ann Arbor, MI, USA, [14]University of Groningen, UMCG, Department of Ophthalmology, Groningen, the Netherlands, [15]Department of Clinical Genetics, Amsterdam University Medical Center (AMC), Amsterdam, the Netherlands, [16]Department of Immunology, Genetics and Pathology, Science for Life Laboratory, Uppsala University, Uppsala, Sweden, [17]University of Groningen, UMCG, Department of Genetics, Groningen, the Netherlands, [18]Estonian Genome Centre, Institute of Genomics, University of Tartu, Tartu, Estonia, [19]Ontario Institute for Cancer Research, Toronto, ON, Canada, [20]Institute for Molecular Medicine Finland, University of Helsinki, Helsinki, Finland, [21]HUNT Research Centre, Department of Public Health and Nursing, NTNU, Norwegian University of Science and Technology, Levanger, Norway, [22]Clinic of Medicine, St. Olavs Hospital, Trondheim University Hospital, Trondheim, Norway, [23]University of Colorado - Anschutz Medical Campus, Aurora, CO, USA, [24]Center for Neuropsychiatric Research, National Health Research Institutes, Miaoli, Taiwan, [25]Bioinformatics Interdepartmental Program, University of California, Los Angeles, Los Angeles, CA, USA, [26]Division of Biostatistics, Institute of Epidemiology and Preventive Medicine, College of Public Health, National Taiwan University, Taiwan, [27]Psychiatric and Neurodevelopmental Genetics Unit, Center for Genomic Medicine, Massachusetts General Hospital, Boston, MA, USA, [28]QIMR Berghofer Medical Research Institute, Brisbane, Australia, [29]Faculty of Health, School of Biomedical Sciences, Queensland University of Technology, Australia, [30]Department of Computer Science, University of California, Los Angeles, Los Angeles, CA, USA, [31]Nuffield Department of Population Health, University of Oxford, Oxford, UK, [32]Department of Epidemiology and Biostatistics, School of Public Health, Peking University Health Science Center, Beijing, China, [33]MRC Population Health Research Unit, University of Oxford, Oxford, UK, [34]Department of Biostatistics and Center for Statistical Genetics, University of Michigan, Ann Arbor, MI, USA, [35]The Charles Bronfman Institute for Personalized Medicine, Icahn School of Medicine at Mount Sinai, New York, NY, USA, [36]deCODE Genetics/Amgen inc., 101, Reykjavik, Iceland, [37]Centre for Genomic and Experimental Medicine, Institute of Genetics and Cancer, University of Edinburgh, Edinburgh, UK, [38]University of Groningen, UMCG, Department of Epidemiology, Groningen, the Netherlands, [39]Department of Anesthesiology, Michigan Medicine, Ann Arbor, MI, USA, [40]Institute of Healthcare Policy & Innovation, University of Michigan, Ann Arbor, MI, USA, [41]Wolfson Institute of Population Health, Queen Mary University of London, London, UK, [42]Chinese Academy of Medical Sciences, Beijing, China, [43]Blizard Institute, Queen Mary University of London, London, UK, [44]Central Pharmaceutical Research Institute, JAPAN TOBACCO INC., Takatsuki 569-1125, Japan, [45]Medical Research Council Human Genetics Unit, Institute of Genetics and Cancer, University of Edinburgh, Edinburgh, UK, [46]Biogen, Cambridge, MA, USA, [47]NIHR Bristol Biomedical Research Centre, Bristol, UK, [48]MRC Epidemiology Unit, University of Cambridge, Cambridge, UK, [49]Center for Precision Psychiatry, Massachusetts General Hospital, Boston, MA, USA, [50]Centre for Population Genomics, VA Boston Healthcare System, Boston, MA, USA, [51]Institute of Precision Health, University of California, Los Angeles, Los Angeles, CA, USA, [52]Program in Neurogenetics,

Department of Neurology, David Geffen School of Medicine, University of California, Los Angeles, Los Angeles, CA, USA, [53]Big Data Institute, Li Ka Shing Centre for Health Information and Discovery, University of Oxford, Oxford, UK, [54]Novo Nordisk Foundation Center for Basic Metabolic Research, Faculty of Medicine and Health Sciences, University of Copenhagen, Copenhagen, Denmark, [55]Department of Computational Biology and Medical Sciences, Graduate school of Frontier Sciences, The University of Tokyo, Tokyo, Japan, [56]University of Michigan, Department of Pediatrics, Ann Arbor MI 48109, [57]School of Basic and Medical Biosciences, Faculty of Life Sciences and Medicine, King's College London, London, UK, [58]Bradford Institute for Health Research, Bradford Teaching Hospitals National Health Service (NHS) Foundation Trust, Bradford, UK, [59]Department of Molecular Genetics, University of Toronto, Toronto, ON, Canada, [60]Department of Human Genetics, David Geffen School of Medicine, University of California, Los Angeles, Los Angeles, CA, USA, [61]Department of Neurology, David Geffen School of Medicine, University of California, Los Angeles, Los Angeles, CA, USA, [62]Institute for Genomic Health, Icahn School of Medicine at Mount Sinai, New York, NY, USA, [63]Department of Public Health & Medical Humanities, School of Medicine, National Yang Ming Chiao Tung University, Taipei, Taiwan, [64]Institute of Behavioral Medicine, College of Medicine, National Cheng Kung University, Tainan, Taiwan, [65]Medical and Population Genomics, Wellcome Sanger Institute, Hinxton, UK, [66]Center for Infectious Disease Education and Research (CiDER), Osaka University, Suita 565-0871, Japan, [67]Laboratory of Statistical Immunology, Immunology Frontier Research Center (WPI-IFReC), Osaka University, Suita 565-0871, Japan, [68]Laboratory for Systems Genetics, RIKEN Center for Integrative Medical Sciences, Yokohama, Japan, [69]Integrated Frontier Research for Medical Science Division, Institute for Open and Transdisciplinary Research Initiatives, Osaka University, Suita 565-0871, Japan, [70]Department of Computational Medicine, David Geffen School of Medicine, University of California, Los Angeles, Los Angeles, CA, USA, [71]Institute for Genetics and Biomedical Research (IRGB), National Research Council (CNR), Cagliari, Italy, [72]Department of Human Genetics, University of Michigan, Ann Arbor, MI, USA
